# Enhancer profiling identifies epigenetic markers of endocrine resistance and reveals therapeutic options for metastatic castration-resistant prostate cancer patients

**DOI:** 10.1101/2023.02.24.23286403

**Authors:** Tesa M. Severson, Yanyun Zhu, Stefan Prekovic, Karianne Schuurman, Holly M. Nguyen, Lisha G. Brown, Sini Hakkola, Yongsoo Kim, Jeroen Kneppers, Simon Linder, Suzan Stelloo, Cor Lieftink, Michiel van der Heijden, Matti Nykter, Vincent van der Noort, Joyce Sanders, Ben Morris, Guido Jenster, Geert JLH van Leenders, Mark Pomerantz, Matthew L. Freedman, Roderick L. Beijersbergen, Alfonso Urbanucci, Lodewyk Wessels, Eva Corey, Wilbert Zwart, Andries M. Bergman

## Abstract

Androgen Receptor (AR) signaling inhibitors, including enzalutamide, are treatment options for patients with metastatic castration-resistant prostate cancer (mCRPC), but resistance inevitably develops. Using metastatic samples from a prospective phase II clinical trial, we epigenetically profiled enhancer/promoter activities with H3K27ac chromatin immunoprecipitation followed by sequencing, before and after AR-targeted therapy. We identified a distinct subset of H3K27ac-differentially marked regions that associated with treatment responsiveness. These data were successfully validated in mCRPC patient-derived xenograft models (PDX). *In silico* analyses revealed HDAC3 as a critical factor that can drive resistance to hormonal interventions, which we validated *in vitro*. Using cell lines and mCRPC PDX tumors *in vitro*, we identified drug-drug synergy between enzalutamide and the pan-HDAC inhibitor vorinostat, providing therapeutic proof-of-concept. These findings demonstrate rationale for new therapeutic strategies using a combination of AR and HDAC inhibitors to improve patient outcome in advanced stages of mCRPC.

## Background

Prostate cancer is the most prevalent cancer type in men, with globally over 1.4 million new diagnoses and 375,000 patients who succumb to the disease each year^1^. Although most patients with high-risk localized disease are effectively treated with either prostatectomy or radiotherapy^2^, eventually 25% of patients will develop metastases for which there is currently no cure^3-5^. Treatment of choice for metastatic prostate cancer patients is androgen deprivation therapy (ADT) which reduces serum testosterone to castration levels, to which virtually all patients initially respond. However, metastatic disease progression despite ongoing ADT, termed metastatic castration-resistant prostate cancer (mCRPC), is inevitable^6^.

Androgen Receptor (AR) is a hormone-dependent transcription factor and the master regulator of prostate cancer development and progression. Upon androgen stimulation, AR alters its conformation, translocates into the nucleus^7^ and associates to the chromatin at distal regulatory elements throughout the genome, hereafter referred to as its ‘cistrome’. AR chromatin binding is facilitated by pioneer factors, such as FOXA1^8^, and operates under tight epigenetic control^8,9^. The majority of active AR sites are hallmarked by acetylation of lysine residue 27 on histone 3 (H3K27ac), a marker of active enhancers and promoters^10,11^. Upon formation of an active transcription complex, AR drives the expression of its target genes to control tumor cell growth. Following progression on ADT, further suppression of the AR signaling axis by new generation AR inhibitors, such as enzalutamide, is an effective treatment for mCRPC patients^12,13^.

Enzalutamide (ENZA) is a well-established therapy for the treatment of mCRPC. It significantly decreases the risk of radiographic progression and death among mCRPC patients in both the pre- and post-chemotherapy settings^12,13^. ENZA blocks AR signaling at multiple levels, including diminished AR chromatin binding, and prevention of coregulator recruitment^14^. However, intrinsic resistance to ENZA is observed in up to 46% of mCRPC patients and duration of response varies greatly between patients^12,13^. Consequently, biomarkers for response prediction to AR-targeted therapeutics, including ENZA, are urgently needed to identify those patients who may benefit from alternative treatment strategies. Moreover, combination treatments to overcome or postpone resistance to AR-targeted therapies, are urgently needed in the clinic.

Several studies have previously compared AR chromatin binding profiles in different disease stages and illustrated plasticity of AR cistromes in tumor development^15,16^ and disease progression^17,18^, being predictive for outcome^17^ and associated with treatment response in cell lines^9^. Despite our expanding knowledge of AR epigenetic alterations in prostate cancer patients and cell line models, there is limited knowledge on FOXA1 cistromics and H3K27ac profiles, in relation to drug resistance in mCRPC patients.

To identify the potential epigenetic alterations that drive ENZA response in mCRPC patients, metastasis-targeted biopsies were collected pre- and post-AR-targeting treatment, while response to treatment was monitored. Chromatin immunoprecipitation followed by massive parallel sequencing (ChIP-seq) was performed on all collected clinical specimens, charting the cistromes of H3K27ac, AR and FOXA1.

Comparative data analyses revealed a specific subset of 657 H3K27ac sites significantly enriched in metastatic lesions from mCRPC patients who did not respond to AR-targeted treatment. These sites were associated with response to castration in mCRPC PDX models, and regulate genes selectively expressed in ENZA-resistant cell line models, illustrating their potential to predict treatment response. Finally, we identified and functionally assessed factors that selectively bind to these 657 resistance-associated H3K27ac sites in cell line models, revealing novel therapeutic candidates and effective drug-drug combinations for treatment-resistant mCRPC patients.

## Methods

### Study design and participants

We conducted a single-arm, open-label, phase 2 study, in patients with mCRPC at the Netherlands Cancer Institute. Male patients over 18 years of age, with histologically confirmed adenocarcinoma of the prostate, Eastern Cooperative Oncology Group (ECOG) performance status 0-2, a serum testosterone level <50 ng/dl, scheduled for ENZA treatment and not previously treated with ENZA, with progressive disease, defined as a PSA rise (PCWG3 criteria^19^) and/or radiographic progression (RECIST 1.1 criteria^20^) and metastatic lesions of which a histological biopsy could safely be obtained, were included in the trial. This single center cohort study was conducted as a sub-investigation of the CPCT-02 biopsy protocol (NCT01855477), which aims to analyze the individual metastatic cancer genome in patients, to develop future personal predictors for response to systemic treatment. Trial procedures, treatment details, patient on-trial monitoring, definition of endpoints, sample size calculations and statistical analysis, are included in the supplementary trial data (Supplementary Data). This study was approved by the local medical ethics committee of the Netherlands Cancer Institute and was activated on January 24^th^, 2012. The protocol complied with the ethical principles of the Declaration of Helsinki. Patients provided signed informed consent for translational studies and recording and analysis of baseline characteristics and clinical outcomes of ENZA treatment.

### Tissue processing and chromatin immunoprecipitation analyses

Biopsies were taken from a lymph-node metastasis or visceral metastasis selected by CT scan, while sites for a biopsy from a bone metastasis were selected by ^68^Ga-PSMA PET scanning. Fresh-frozen metastatic biopsy samples from 64 CRPC patients was collected. The tumor percentage of these samples was scored on hematoxylin and eosin (H&E) stained slides by a dedicated pathologist. After tumor cell content was confirmed, chromatin immunoprecipitations were performed for AR, FOXA1 and H3K27ac at qualified samples as previously described^21-23^. In brief, lymph-node and visceral samples were cryo-sectioned into slices of 30μm, while bone samples were cryo-sectioned to slices of 10μm, and crosslinked using DSG (20593; Thermo Fisher Scientific) for 25 min. For each sample, 5μg of antibody and 50μl of either Protein A or Protein G magnetic beads (10008D or 10009D; Thermo Fisher Scientific) were used. Inputs for individual patients were generated as controls. Antibodies used were: AR (Millipore, 06-680), FOXA1 (Abcam, ab5089) and H3K27ac (Active motif, 39133). Libraries were prepared and sequenced using the Illumina HiSeq2500 (65 bp, single end).

### ChIP-seq data analysis

Raw sequence data were aligned to hg19 using BWA v0.5.20. Aligned reads were filtered for mapping quality (MQ) > 20 using samtools v1.8^24^. Duplicate reads were marked using Picard MarkDupes function v2.18 (http://broadinstitute.github.io/picard/). Peaks were called using macs2 (v2.1.1)^25^ with the fragment size determined using Phantompeakqualtools^26^ against corresponding input DNA for all samples. Phantompeakqualtools was used to identify the Relative Strand Cross-correlation (RSC)^26^ and deepTools (v2.0)^27^ to determine the fraction of reads in peaks (FRiP) and readcounts. Samples with RSC > 0.7, FRiP ≥ 1.0 and ≥ 8,000 peaks were kept for further analysis (Supplementary Table 3). Snapshots of raw signal were generated using pyGenomeTracks (v3.6) with bigwig files^28^. Bigwig files were generated from aligned bam files using deepTools v2.0 bamCoverage function. To correlate read count data in 50kb bins across the genome for all samples and PDX samples, deepTools computeMatrix function was used on bigwigs followed by plotCorrelation. Visualization of raw reads was carried out with bigwigs using deepTools (v2.0) computeMatrix, plotHeatmap and plotProfile functions. For visualizing profiles of binding data between groups (non-responders/responders) at specific regions, aligned files from the samples within groups were merged and subsequently downsampled to equivalent readcounts (∼20 million reads) using samtools and visualized using deepTools plotProfile. Principal component analysis was carried out using plotPCA function with the reads counted in peaks (dba.count function) from DiffBind package v2.4.8 in R v3.4.4. Supervised differential analyses using dba.analyze and resulting heatmap and volcano plot were generated using the DiffBind package (v2.4.8 in R v3.4.4) with the reads counted in peaks using the dba.count function using default DESeq2 method. Volcano plot hexbin density tiles were plotted with R package hexbin (v1.28.1). Genomic features were assigned to differential peaksets using ChIPSeeker (v1.26.2)^29^ in R (v4.0.3).

To compare H3K27ac signal across various samples in our non-responder H3K27ac regions of interest, we first downloaded public PDX mCRPC H3K27ac ChIP-seq data^18^ (GSE130408) and additional H3K27ac data from primary prostate cancer patient tumors (e.g. Gleason 7,9, cases, controls (GSE120738))^30^ and aligned as above. In addition, we examined H3K27ac data in the same manner from in-house generated datasets from treatment naïve metastatic samples, non-responder metastatic samples (this study), and publicly available healthy and primary tumor tissue (GSE130408)^18^. Visualizations of these data were generated with deepTools v2.0 computeMatrix followed by plotHeatmap of individual files and plotProfile as described above for binding data between groups.

For gene set enrichment, genes associated with H3K27ac non-responder sites (within 50kb of a transcription start site (TSS)) were identified in R (v4.0.3) using the ChIPSeeker package (v1.26.2)^29^. Enrichment tests for gene sets were performed in R (v4.1.2) using the GeneOverlap package (v1.30.0) and visualized in ggplot (v3.4.0). The average gene expression of these genes was calculated and plotted in public scRNA-seq from parental LNCaP cells and cells exposed to ENZA until resistance arose (RES-B^31,32^) using the Seurat package (v4.3.0)^33^ in R (v4.1.2).

Readcounts and fraction of reads in peaks (FrIP) were visualized using the ggplot2 v2.3.3.0 with Wilcoxon tests performed with ggpubr package v0.3.0 in R v3.5.0. To determine significant enrichment of our intervals with publicly available ChIP-seq data we queried our intervals against the CistromeDB transcription factor dataset^34^ using the GIGGLE search function^35^. Prostate and prostate cancer experiments (Supplemental Table 5) were selected specifically for analysis. A scatterplot of the mean Enrichment score (combo_score) for each factor was generated using ggplot v2.3.3.0 in R v3.5.0 (Supplemental Table 5).

### Patient-derived xenograft studies

All animal experiments were performed after University of Washington IACUC approval following ARRIVE and NIH guidelines. Subcutaneous tumors were implanted in intact C.B. 17 SCID male mice (Charles River) and when tumors reached 100 mm^3^ animals were randomized to control and castrated groups. Tumor growth and body weights were monitored twice a week. Animals were sacrificed at the end of the study or when animals became compromised. Responses to castration were also fully described previously^36^. The doubling time was estimated using exponential (Malthusian) growth model. If only one value was available, doubling time was not computed. For samples with a negative doubling time, the value was re-normalized to the mean value of the corresponding control model yielding a positive value. Significant differences between classes determined by one-way ANOVA followed by post-hoc Tukey HSD test. Visualization was carried out in R using ggplot (v3.3.6)

### Cell lines and culture conditions

Castration-resistant prostate cancer models (LNCaP-Abl and LNCaP-16D^37^) were kindly provided by Helmut Klocker^38^ and Amina Zoubeidi^39^. ENZA resistant LNCaP derivatives LNCaP-Enz^R^ were kindly provided by the Donald Vander Griend^40^. LNCaP-Abl cells were cultured in RPMI-1640 medium containing 10% DCC (hormone deprived FBS), LNCaP-16D^37^ cells were cultured in RPMI-1640 medium supplemented with 10% FBS, and LNCaP-Enz^R^ cells were cultured in RPMI-1640 medium containing 10% FBS and 10μM ENZA. All cell lines were authenticated and tested for mycoplasma contamination.

### siRNA screen proliferation assay and analyses

siRNAs were purchased from Dharmacon (Lafayette, CO, USA). Non-targeting siRNA and siPLK1 were applied as positive and negative controls. 5μl of 50nM siRNA pools were seeded in individual wells of a 96 well-plate. Cells were reverse transfected with 5μl 1% Lipofectamine RNAiMAX Reagent (Invitrogen, Eindhoven, Netherlands) in Optimem (Thermofisher, Eindhoven, Netherlands) in 90μl culture medium. For LNCaP-Abl and LNCaP-16D, 10.000 cells were seeded per well, and 20.000 cells for LNCaP-Enz^R^ cells. Optimal experimental setup was determined for each cell line and after 7 (LNCaP-16D), 9 (LNCaP-Abl) and 10 (LNCaP-Enz^R^) days, cell viability was determined using CellTiter-Blue and values were normalized over siControl. After incubating for 3 hours, viability was measured using a fluorescence reader (EnVision 2014).

The primary pooled siRNA and validation deconvolution screen were analysed in the following way. Using the CellTiter-Blue measurements of the positive and negative controls, a z’factor was calculated per plate and plates with a z’factor < 0 were removed from the dataset. The data was then normalized using Normalized percent inhibition^41^. After normalization, correlations between replicate plates were calculated and plates which did not correlate well with the other replicate plates, were removed. Over the replicates a mean value was calculated. Per condition a normalized distribution for mean values of the negative controls was approximated based on mean and SD value, and used to calculate for each pooled siRNA a p-value, which was corrected for multiple testing using the Benjamini-Hochberg method. From the primary screen an initial selection was made of the siRNA pools that were a hit in at least two out of three cell lines, which produced a list of 11 hits. The 11 hits from the primary pooled screen were subsequently selected for a deconvolution validation screen, in which four individual siRNAs were tested separately. All targets with two individual siRNAs with among replicates a mean ≤ 0.7 and FDR ≤ 0.1, where considered validated hits. All calculation were done in R.

Expression levels per target gene in siRNA deconvolution experiments were assessed by means of qPCR analysis, using specific primer-pairs for ACTB (5’-CCTGGCACCCAGCACAAT-3’, 5’-GGGCCGGACTCGTCATACT-3’), FOXA1 (FW 5’-GTGAAGATGGAAGGGCATGAA-3’, REV 5’-CCTGAGTTCATGTTGCTGACC-3’), ASH2L (FW 5’-CTGACGTCTTGTATCACGTG-3’, REV 5’-GCATCTTTGGGAGAACATTTG-3’), GATA2 (FW 5’-GACAAGGACGGCGTCAAGTA-3’, REV 5’-GGTGCCCATAGTAGCTAGGC-3’) and HDAC3 (FW 5’-ACGGTGTCCTTCCACAAATACG-3’, REV 5’-GGTGCTTGTAACTCTGGTCATC-3’). In brief, after siRNA transfection using the abovementioned protocol RNA was isolated using RNAGEM kit (MicroGEM), and quantified by Quant-iT™ RiboGreen™ RNA Assay Kit (Thermo Fisher Scientific), following quantification cDNA was synthesized using the SuperScript™ III Reverse Transcriptase system (Thermo Fisher Scientific, USA) with random hexamer primers according to the instructions provided by manufacturers. Quantitative PCR (qPCR) was performed using the SensiMix™ SYBR Kit (Bioline, UK) according to the manufacturer’s instructions on a QuantStudio™ 6 Flex System (Thermo Fisher Scientific, USA). All data was firstly normalized over ACTB expression, and then over the siControl values. For all primer pairs, 2 biological replicates with 2 technical replicates each were analyzed.

### PDX in vitro studies

Subcutaneous tumors were harvested and dissociated using the Miltenyi gentleMACS system with a Human Tumor dissociation kit (Miltenyi Corp). Cells were seeded in clear bottom white -walled flat bottom 96-well plates (20,000 cells per well) in RPMI and 10% FBS. ENZA (MedChem Express) and vorinostat (MedChem Express) 10mM in DMSO and diluted with RPMI to indicated concentrations. Effects of the treatments were evaluated after 5 days using CellTiter-Glo (Promega).

### Drug synergy assessment

In a 384-well plate, 500 LNCaP or LNCaP-16D cells were seeded and treated with various concentrations of ENZA (MedChemExpress, Monmouth Junction NJ, USA) and vorinostat (kindly provided by Rene Bernards, NKI). Five days later, the CellTiter Glo assay (Promega Benelux BV, Leiden, Netherlands) was performed according to the manufacturer’s instruction. All the assays were performed in biological quadruplicates (n=4). All the conditions (single and combination) were normalized to non-treated condition (set at 100). SynergyFinder 2.0^42^ was used to evaluate and plot synergistic potential using highest-single agent (HSA) synergy reference model. Response of the two cell lines to single agent vorinostat was also investigated and plotted using the normalized viability in full media (FBS) and area under the curve (AUC) method.

### Single-cell analyses

Single-cell RNA-seq (GSE168669) data was used to produce UMAP visualizations of LNCaP parental and LNCaP RES-B retaining cluster identities from Taavitsainen *et al*.^32^ The genes proximal to the 657 non-responder H3K27ac sites were compiled into a gene set expression analysis to produce scores per cell with AddModuleScore function from Seurat (version 4.3.0). Previously identified single-cell clusters (clusters 0 to 12)^32^ were used in the enrichment analysis to overlap genes proximal to the 657 non-responder H3K27ac sites.

## Results

### Phase II trial of AR-targeted therapy in patients with mCRPC

To identify novel epigenetic biomarkers, we conducted a single-arm, open-label, phase 2 study in patients with mCRPC treated with a new line of AR targeted therapy, being a sub-investigation of the CPCT-02 biopsy protocol (NCT01855477) (Figure 1A). Between September 2014 and April 2019, a total of 64 mCRPC patients were enrolled in the trial. Baseline characteristics are summarized in Supplementary Table 1, trial outcomes in Supplementary Table 2 and Supplementary Figure 1: Consort diagram, with in-depth description of relevant clinicopathological parameters and outcomes included in the supplementary clinical trial data section (Supplementary Data).

**Figure 1:**
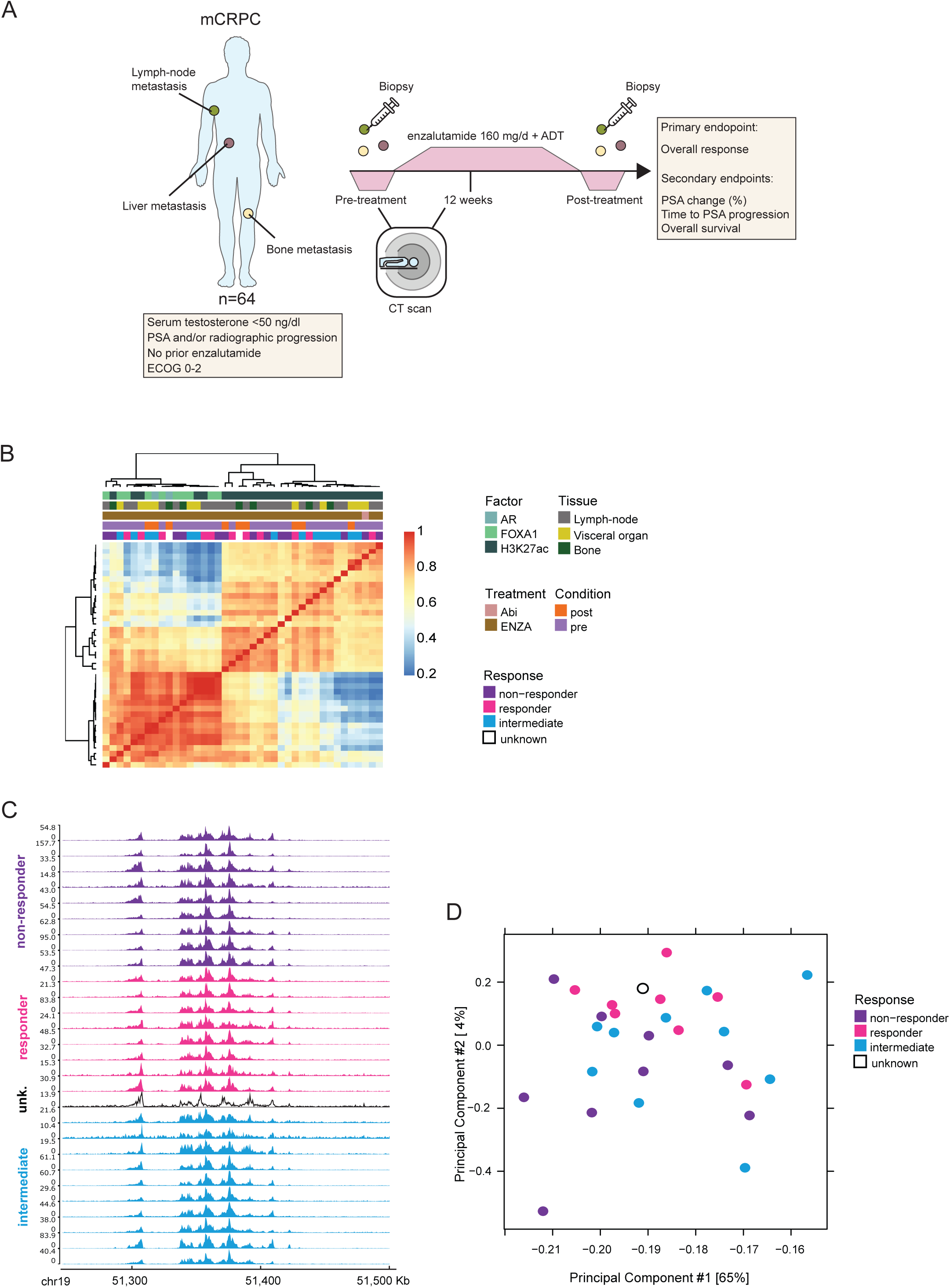
Clinical trial design and ChIP-seq data collection. A. Setup of the clinical trial. Patients with mCRPC are enrolled in the study, and an imaging-guided biopsy is taken prior to onset of enzalutamide treatment. One patient in the study was treated with abiraterone. B. Correlation heatmap of ChIP-seq data (50kb bins across the genome, Pearson correlation) for H3K27ac, AR and FOXA1 among all mCRPC samples (n=40). Colors bars indicate ChIP factors: AR (light blue), FOXA1 (light green) and H3K27ac (dark green), tissue of sample origin: lymph-node (grey), visceral organ (yellow) and bone (dark green), treatment: abiraterone (Abi), salmon)) and enzalutamide (ENZA), brown)), condition of the sample: pre-treatment (purple) and post-treatment (orange), and treatment response: non-responder (dark purple), responder (pink), intermediate (blue) and unknown (black outline). C. Snapshot of H3K27ac ChIP-seq (n=28) in different treatment response groups: responders (pink), non-responders (purple), unknown ((unk.), black outline) and intermediate (blue). The read counts (left) and genomic coordinates (bottom) are indicated. D. Principal Component Analysis using normalized read counts in all peaks in H3K27ac ChIP-seq data (n=73039) from all samples (n=28). Samples labeled according to responders (pink), non-responders (purple), intermediate (blue) or unknown (white).

### Biopsy assessment and evaluable population for biomarker discovery

All 64 patients had a pre-treatment biopsy from a metastatic lesion. Biopsy sites from the whole population include bone (n=19; 29.7%), lymph-nodes (n=32; 50.0%) and visceral organs (n=13; 20.3%) (Supplementary Table 1). A second biopsy (post-treatment) was taken upon disease progression for 15 patients. Biopsies with ≥30% tumor cells were further processed for downstream molecular analyses (42 and 12 for pre- and post-treatment, respectively) (Supplementary Figure 1: Consort diagram). We successfully generated ChIP-seq data for active promoter/enhancer histone modification H3K27ac —passing stringent QC requirements (See Methods) for 22 out of the 42 samples. Four (18.2%) of the pre-treatment biopsies evaluable for biomarkers, were from bone metastases, while 13 (59.1%) were from lymph-node metastases and 5 (22.7%) from visceral metastases (Supplementary Table 1). For 6 of 15 post-treatment biopsies, we obtained high quality H3K27ac ChIP-seq data. One (16.7%) of the post-treatment biopsies was from bone, 3 (50.0%) were from lymph-nodes and 2 (33.3%) were from visceral organs. One patient had both pre- and post-treatment biopsies resulting in 28 biopsy samples for further analyses from 27 unique patients. The baseline characteristics and treatment outcomes of the 22 patients who donated an evaluable pre-treatment biopsy and the 6 patients who donated an evaluable post-treatment biopsy, are summarized in Supplementary Table 1 and Supplementary Table 2, respectively. There were no significant differences in baseline age, serum PSA, the treatment outcomes, duration of treatment, PSA change from baseline, and Time to PSA Progression (TTPP) between the patients who donated an evaluable pre- and/or post-treatment biopsy and the whole population (Supplementary Table 1 and Supplementary Table 2). Additional clinical information and methods can be found in Supplementary Information.

Prior to functional genomic downstream analyses, all patients were categorized for their overall response, which was a composite of three outcomes: (1) ≥ 50% PSA decrease from baseline, (2) radiographic response (stable disease, partial response or complete response), and (3) longer than median TTPP. Assessment was conservative; in case a patient could not be evaluated on a particular outcome measure, it was considered as no response. All patients were evaluable for this endpoint, except for one patient (1.6%) who could not be evaluated for any of the three outcomes (Supplementary Table 2). In the whole population, 23.4% of patients scored on all three items (Response to ENZA), 37.5% of patients did not score on any item (No response to ENZA) and the remaining 37.5% of patients had inconsistent responses on the three outcome measures listed above (Intermediate response to ENZA). Of the 22 patients in the pre-treatment evaluable population, 6 (27.3%) patients had a response, 8 (36.4%) patients had an intermediate response, and 8 (36.4%) patients had no response to ENZA (Supplementary Table S2). Of the 6 patients in the post-treatment evaluable population, 2 patients had a response (33.3%), 2 patients had an intermediate response (33.3%), 1 patient had no response (16.7%) to ENZA treatment, while 1 patient was not evaluable (16.7%) (Supplementary Table S2). Based on the ChIP-seq QC parameters and clinical assessment of our trial data, 28 biopsies (22 pre- and 6 post-treatment, from 27 unique patients) with high quality ChIP-seq data are available, roughly equally sized response groups were formed, with 8 responders, 10 intermediate and 9 non-responders to treatment (one unknown) (Figure 1B).

### Genome-wide epigenetic profiling of mCRPC

Apart from H3K27ac ChIP-seq data, for which we successfully generated high-quality data for 28 metastatic biopsy samples (see above), 10 FOXA1 ChIP-seq and 2 AR ChIP-seq datasets were generated on these fine needle core biopsies (Figure 1B and Supplementary Figure 1: CONSORT diagram). As all these patients received prior ADT, the low circulating testosterone levels may explain the relatively low success-rate of AR ChIP-seq (of which chromatin binding is decreased following ADT) as compared to FOXA1. For peak numbers, read counts and other relevant ChIP-seq QC parameters, see Supplementary Table 3 and Supplementary Figure 2A,B.

All H3K27ac ChIP-seq samples were highly correlated based on genome-wide patterns (Figure 1B) indicating low inter-tumor heterogeneity, and robust technical reproducibility. As expected, and in line with our previous study on multiple metastases from the same patient^22^, FOXA1 and AR profiles were intermingled in our unsupervised hierarchical analysis reflecting the direct biological interplay between these two factors^16,43^. No correlation was observed with metastatic site or treatment condition/status or clinical response in the clustering with all factors (Figure 1B), nor on H3K27ac alone (Supplementary Figure 3A,B). As H3K27ac ChIP-seq data represented the largest and most-complete dataset, we decided to first focus on these samples. For H3K27ac profiles, we found comparably high-quality peaks across all three response groups, as exemplified on single locus (Figure 1C) and genome-wide scale (Figure 1D). Taken together, our data indicate the vast majority of H3K27ac sites are overlapping across sample types, irrespectively of AR-targeted therapy response.

### Distinct H3K27ac profiles identify mCRPC tumors resistant to AR-targeted therapy

While the total universe of H3K27ac profiles did not differ between response groups of patients, possible subsets of regions may still exist that stratify patients on outcome. To ensure that subtle differences in cut-offs and definitions of treatment response would not affect data interpretation, we performed supervised differential binding analysis^44^ with the H3K27ac data in the most extreme treatment response groups (responder and non-responder) (Figure 2A). In total, we observed 682 H3K27ac regions that significantly differed between these response groups, with 657 sites selectively enriched in non-responder patients and merely 25 sites found selectively enriched in responders (adj.p ≤ 0.05, logFC ≥ abs|2|, Figure 2B). As expected for H3K27ac ChIP-seq, both sets of sites are predominantly found in distal intergenic regions (Supplementary Table 4). Differentially enriched peaks between responders and non-responders were robust, as exemplified for three genomic loci (Figure 2C), and quantified showing enriched signal in non-responders compared to responders across all non-responder sites (Figure 2D).

**Figure 2:**
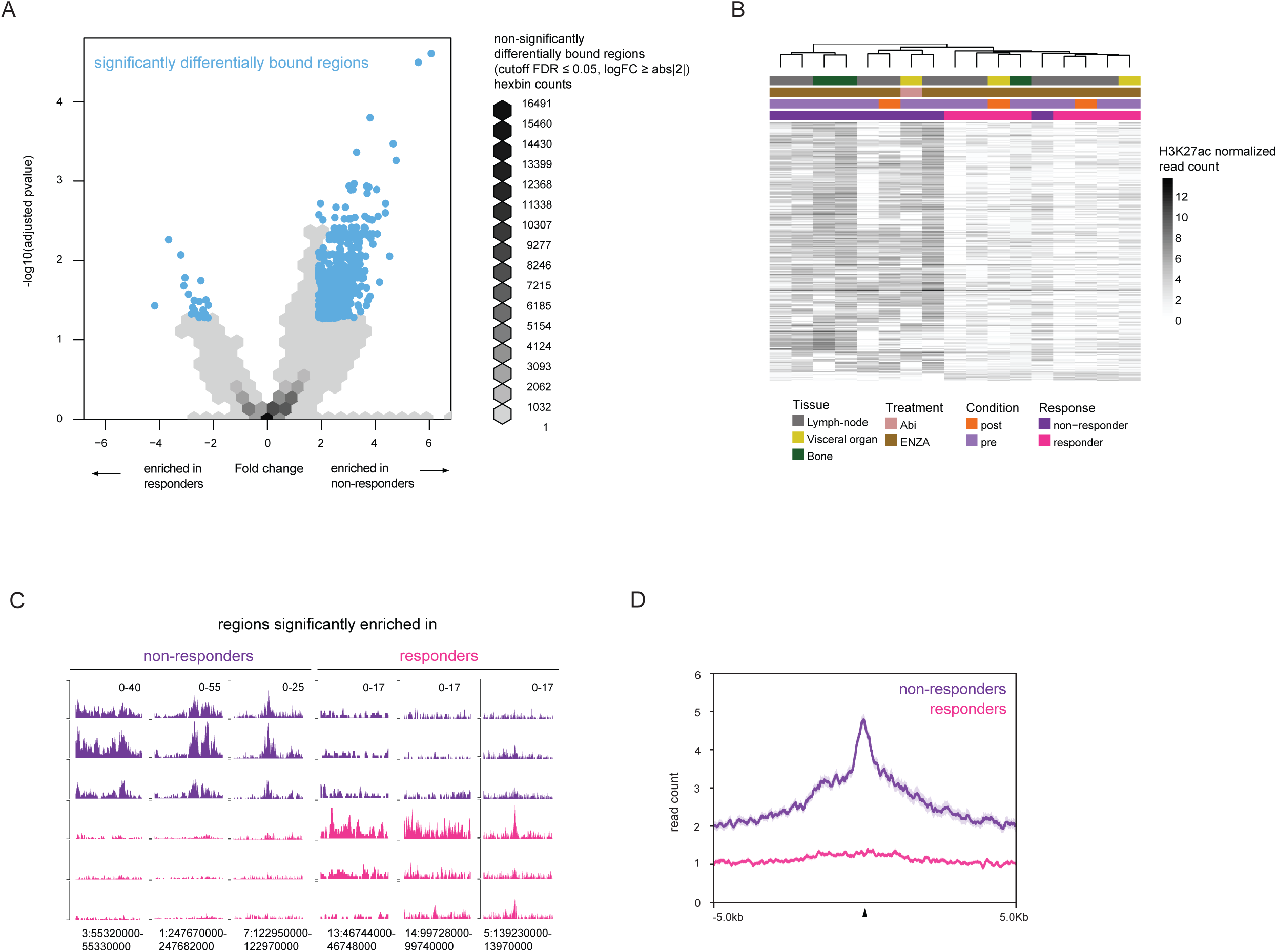
Distinct H3K27ac profiles stratify mCRPC patients on response to AR inhibition. A. Differentially enriched regions from H3K27ac ChIP-seq data visualized by volcano plot (n=73039). Regions marked by blue dots were significant (DiffBind DESeq2 two-tailed adjusted p-value ≤ 0.05, logFC ≥ abs|2|) (n=848); all other regions are shown with hexbin density to avoid over-plotting (n=72191). Each data point density tile (hexagon) represents density of data within the tile from low (light grey) to high (black). B. Heatmap showing normalized read count of H3K27ac data in significantly differentially bound regions (DiffBind DESeq2 two-tailed adjusted p-value ≤ 0.05, logFC ≥ abs|2|) (n=848) in responder (n=8) and non-responder samples (n=9). Colors bars indicate tissue of sample origin: lymph-node (grey), visceral organ (yellow) and bone (dark green), treatment: abiraterone (Abi), salmon)) and enzalutamide (ENZA), brown)), condition of the sample: pre-treatment (purple) and post-treatment (orange) and treatment response: non-responder (dark purple) and responder (pink). C. Individual snapshot of H3K27ac enriched differently in 3 responder patients (pink) and 3 non-responder patients (purple) as examples. The read counts and genomic coordinates are indicated (top right and bottom, respectively). D. Average H3K27ac read count profiles of all merged data for responder patients (pink, n=8) and all merged non-responder patients (purple, n=9) at the 657 non-responder enriched sites (±5 kb from the peak center). Shading indicates standard-error of the data.

To determine whether the 657 H3K27ac sites enriched in non-responders represent an acquired feature of mCRPC, or whether H3K27ac signal at these regions is already present in the primary disease setting and associated with aggressiveness, we re-analyzed H3K27ac ChIP-seq data from a matched case-control cohort of treatment naïve primary prostate cancer patients that we reported previously^30^. We observed no difference in H3K27ac signal at these sites based on case/control status (Supplementary Figure 4A), nor on Gleason score (7 versus 9) (Supplementary Figure 4B) while typical signal for known primary-specific AR binding sites was clearly present^16^ (Supplementary Figure 4C). Further supporting the notion that these regions are acquired in the treatment-resistance metastatic setting, we observed stronger signal in the non-responder metastatic samples (this study) compared with treatment naïve metastasis samples and previously reported primary prostate cancer samples^18^ as well as healthy prostate tissue^18^ (Supplementary Figure 4D). Together, these data suggest that the resistance-associated H3K27ac sites represent an mCRPC-unique feature of resistance to AR-targeted therapy. Collectively, our data indicate that a specific subset of H3K27ac sites enables us to stratify mCRPC patients for an outcome to third generation AR inhibitor treatment.

### Resistance-associated H3K27ac profiles predict response to castration in mCRPC patient-derived xenografts

H3K27ac profiling in clinical samples allowed us to stratify tumors from mCRPC patients on response to AR-targeted therapeutics. To independently validate these findings and to explore the potential for stratification beyond our own study, we next investigated an existing H3K27ac ChIP-seq dataset that we previously reported for mCRPC PDX samples^18^. The PDX models were generated from CRPC prostate cancer tumors, and represent metastatic samples from multiple metastatic sites, including adrenal glands, ascites, bladder, bone, bowel, lymph-node and liver^36^. Originally, to determine the hormone-dependency of the PDX tumor growth, PDX tumors were grown in testosterone proficient male mice, after which the animals were either castrated or left intact (see overview in Figure 3A).

**Figure 3:**
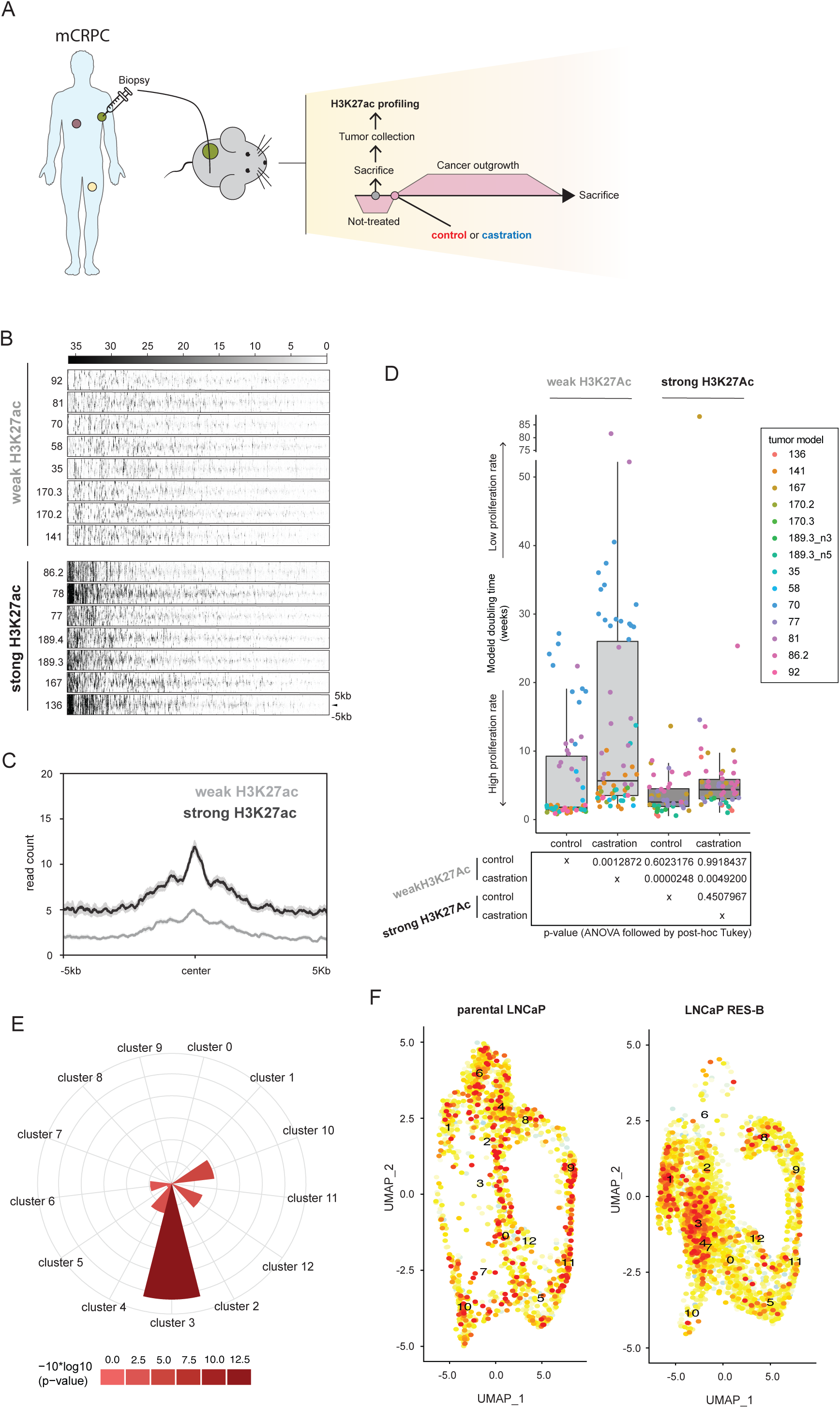
mCRPC PDX validations of resistance-associated H2K27ac regions. A. Overview of the PDX models setup. Prostate cancer samples from mCRPC patients were obtained and implanted into the mouse to establish PDXs. These PDXs were characterized previously with their response to castration by the change of tumor volume. B. Heatmap depicting raw read counts of H3K27ac ChIP-seq signal from PDX samples at the non-responder enriched 657 H3K27ac regions, identified from the mCRPC patient samples (±5 kb from the peak center). C. Average H3K27ac read count profiles of all PDX merged data for samples with weak (grey, n=8) and strong (black, n=7) signal in the non-responder 657 H3K27ac regions. (±5 kb from the peak center). Shading indicates standard-error of the data. D. Box plots depicting doubling time of PDX models estimated using exponential (Malthusian) growth model (y-axis) by group (x-axis). The central mark indicates the median, and the bottom and top edges of the box indicate the 25^th^ and 75^th^ percentiles, respectively. The maximum whisker lengths are specified as 1.5 times the interquartile range. All individual values are depicted as circles colored by PDX model (strong H3K27ac – Castration: n=57, strong H3K27ac – Control: n=45, weak H3K27ac – Castration: n=71, weak H3K27ac – Control: n=73). Table below indicates the p-values obtained for one-way ANOVA followed by Tukey HSD test for all combinations. E. Polar plot reporting the -10*log_10_(p-values) (Fisher’s exact test) of gene overlap enrichment tests between genes associated with H3K27ac non-responder regions and LNCaP scRNA-seq cluster marker genes. Color indicates strength of significance from low (pink) to high (red). F. Uniform manifold approximation and projection (UMAP) visualization showing the average gene expression score of genes associated with H3K27ac non-responder regions in the parental LNCaP (left) and the ENZA-Resistant, RES-B (right) single cells. Original scRNA-seq clusters (0-12) are superimposed on each plot.

**Figure 4:**
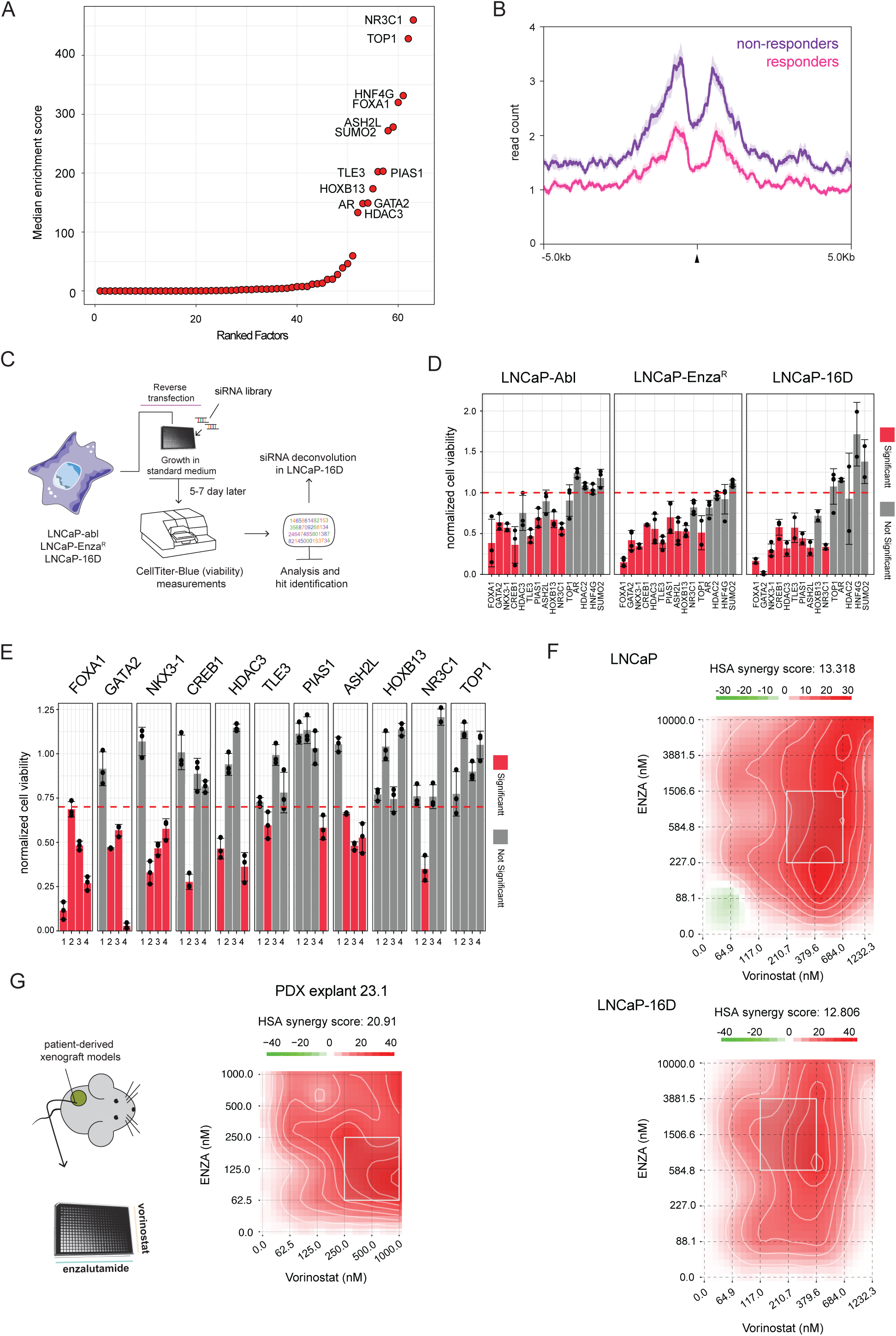
Characterization of the non-responder enriched H3K27ac sites reveals drivers of resistance. A. Enrichment analysis to determine significant overlap of 657 H3K27ac non-responder sites with publicly available ChIP-seq data for factors previously studied in prostate cancer cell lines (n=863). Graph shows median enrichment score for each factor (GIGGLE combo score, indicating low to high significant enrichment score (Fisher’s exact two-tailed test and odds-ratio)). Factors are ordered by highest score (enrichment) in the dataset with text shown in those with median enrichment score > 100. B. Average FOXA1 read count profiles of merged data, at the 657 non-responder enriched H3K27ac sites (±5 kb from the peak center), comparing responders (pink, n=8) and non-responders (purple, n=9). Shading indicates standard-error of the data. C. Setup of siRNA screen to identify factors critical of prostate cancer cell line viability, resistant to androgen ablation or enzalutamide treatment. D. Screen results for pooled siRNAs, showing decreased viability of prostate cancer cell line models LNCaP-Abl (left), LNCaP-Enz^R^ (middle) and LNCaP-16D (right). Cell viability was determined by CellTiter-Blue, and data are normalized over siControl. Bars indicate mean values ± SD (n≥2). Adjusted P values (padj) were determined by two-sided t-test with multiple testing correction (Benjamini-Hochberg method). Statistically significant conditions (padj <0.05) are shown in red. E. siRNA deconvolution experiment, separately analyzing each individual siRNA for the 11 remaining hits in LNCaP-16D cells on cell viability. Cell viability was determined by CellTiter-Blue and data are normalized over siControl. Bars indicate mean values ± SD (n≥3). Adjusted P values determined as above. Statistically significant conditions (padj <0.05) are shown in red. F. Enzalutamide-vorinostat drug synergy analyses for LNCaP (top) and LNCaP-16D (bottom) based on viability experiments performed after 5 days of treatment. Vorinostat and enzalutamide (ENZA) concentrations in nM on the x- and y-axis, respectively. Synergy determined using the HSA model (score >10 indicates synergy with regions of maximal synergy outlined in white). Graphs representative for 4 biological replicates. G. (Left) Experimental setup of tumor explant studies. Tumor samples are removed from the animals, and exposed *ex vivo* to increasing concentrations of enzalutamide and vorinostat and assessed for viability. (Right) Drug synergy representation for mCRPC PDX explant LuCaP 23.1. Synergy determined using the HSA model (score > 10 indicates synergy with regions of maximal synergy outlined in white).

To evaluate whether the PDX tumors would classify according to the 657 non-responder H3K27ac sites, we plotted H3K27ac ChIP-seq data for these sites in 15 available PDX samples (Figure 3B and Figure 3C). Interestingly, 7 PDXs (45%) displayed strong H3K27ac signal at these regions, while the remaining 8 PDX samples (55%) displayed weak signal at these sites; with no significant global differences between these two groups of samples (Supplementary Figure 5). Integrating these cistromic data with the *in vivo* response-to-castration data showed that a strong H3K27ac signal is correlated with less tumor regression upon castration of the animals (Figure 3D, raw data Supplementary Figure 6). In contrast, PDXs with weak H3K27ac signal at our clinically observed H3K27ac non-responder sites, showed tumor regression upon animal castration. Importantly, using single-cell RNA-seq data (scRNA-seq) from prostate cancer cell lines, we find genes associated with our clinically observed H3K27ac non-responder sites as significantly enriched in a cell cluster that selectively appears in LNCaP-derived ENZA-resistant cells^32^ (cluster 3) (Figure 3E). After long-term treatment with ENZA, cluster 3 enriched mainly for LNCaP RES-B resistant cells and only partially for RES-A cells. In fact, in LNCaP RES-B cells cluster 3 was expanded compared to parental cells and also compared to RES-A, indicating that these cells drive resistance-specific biology. In addition, average gene expression of genes associated with H3K27ac non-responder sites was linked with cluster 3 expansion (Figure 3F). Interestingly, scATAC-seq (single cell Assay for Transposase-Accessible Chromatin using sequencing) clusters identified previously to be associated with scRNA-seq cluster 3^32^, indicated no selective enrichment of known transcription factors involved in treatment-mediated chromatin reprogramming in prostate cancer. Together, these data illustrate that distinct H3K27ac signals stratifying patients for response to AR blockade, and we validated the same profiles in mCRPC PDX models and at the single-cell level in models of ENZA resistance.

### Driver identification for resistance to AR inhibition in mCRPC

Using the combined datastreams of H3K27ac ChIP-seq and response-to-castration in PDX models, we successfully validated our 657 H3K27ac sites as indicative for unresponsiveness to hormonal intervention in mCRPC. As no selective transcription factor usage was enriched upon integrating scRNA-seq and scATAC-seq data, we conclude that possibly multiple factors -or proteins binding the genome without direct DNA recognition motif, drive resistance. Therefore, instead of TF motif analyses, we used GIGGLE; a genomics search engine that queries previously-reported protein/chromatin occupancy datasets and ranks the significance of genomic loci shared between query and a database of regions^35^. Specifically, we analyzed an extensive database of ChIP-seq experiments from prostate tissue-derived cell lines and prostate cancer cell lines^34,45^ (Supplementary Table 5) to explore which DNA-associated proteins bind at our defined non-responder H3K27ac sites (Figure 4A). This analysis identified multiple factors previously reported to drive resistance to ENZA treatment or castration, including HNF4G^46^, NR3C1 (glucocorticoid receptor)^47-49^ and FOXA1^50^ (Supplementary Table 5). To clinically validate the cell line-based GIGGLE enrichment data, we next analyzed the FOXA1 ChIP-seq data from our mCRPC samples, separating the non-responder (n=4) and responder (n=3) samples. These analyses revealed selective enrichment of FOXA1 binding at the 657 non-responder H3K27ac sites in mCRPC samples from non-responder patients, confirming the GIGGLE enrichment data (Figure 4B).

Next, we sought to explore the functional involvement of top-enriched factors (Supplementary Table 5) in driving resistance to AR blockade (essential genes in our setting), by designing and performing a focused siRNA screen (4 pooled siRNAs per target (genes with median GIGGLE combo enrichment score >20)) to target genes in two cell line models of castration resistance (LNCaP-Abl^38^ and LNCaP-16D^37^) as well as a model of ENZA resistance (LNCaP-Enz^R^)^40^ (Figure 4C). From the pooled siRNA experiments, eleven hits that significantly diminished proliferation in at least two out of three cell lines (Figure 4D), relative to siControl and were identified as top-enriched factors in the GIGGLE analysis, were selected for deconvolution experiments in castration-resistant LNCaP-16D. Single siRNAs were tested individually, in which decreased cell proliferation potential observed for at least 2 out of 4 siRNAs was considered a validated hit. These analyses identified factors previously described as critical in driving resistance to both ENZA and castration in prostate cancer cell lines: FOXA1^50^ and GATA2^51^ (Figure 4E, Supplementary Figure 7). Furthermore, two factors previously reported to be associated with castration resistance, but not studied before for their potential involvement in driving resistance to ENZA, were identified: HDAC3^52^ and ASH2L^53^. Collectively, these studies revealed potential drivers and possible drug targets to treat castration-resistant prostate cancer.

### Histone deacetylase (HDAC) inhibition synergizes with enzalutamide to block mCRPC cell growth

Computational analyses and perturbation studies identified five factors of potential therapeutic interest. As transcription factors are considered challenging drug targets, we focused on HDAC3 for further downstream studies. HDAC3 has previously been reported as therapeutic target in castration-resistant prostate cancer^52^, but remains unexplored in the ENZA-resistant setting. Highly selective HDAC3 inhibitors have been described but have not been explored for efficacy and tolerability in clinical trials^54^. Less specific HDAC inhibitors are well characterized and clinically implicated in the treatment of several cancer types, including vorinostat in the treatment of cutaneous T-cell lymphoma^55^. Vorinostat has been previously reported to block proliferation of prostate cancer cells^56^ and to synergize with the AR-antagonist bicalutamide^57^. Consequently, HDAC inhibitors have the potential to overcome resistance to established mCRPC treatments, including AR targeted drugs^58^. A significant increase in sensitivity to HDAC inhibition was observed in castration-resistant LNCaP-16D cells relative to hormone-sensitive parental LNCaP cells (Supplementary Figure 8. Importantly, in both LNCaP cells and LNCaP-16D cells, vorinostat synergized with ENZA (Figure 4F, Supplementary Figure 9A). To further establish therapeutic proof-of-concept, subcutaneous PDX tumors were dissociated and treated *ex vivo* with increasing concentrations of ENZA, vorinostat, or both, which allowed us to determine synergy (Figure 4G). In agreement with the cell line-based results, *ex vivo* drug response in mCRPC PDXs confirmed synergistic interactions between vorinostat and ENZA (Figure 4G, Supplementary Figure 9A, 9B).

In summary, by performing H3K27ac ChIP-seq analyses of metastatic lesions from mCRPC patients, we identified an epigenetics-based classification for response prediction to AR-targeted therapy which we successfully validated in mCRPC PDX mouse models. These analyses revealed drivers for resistance to AR-targeted therapeutics, and identified ENZA in combination with pan-HDAC inhibitor vorinostat as a novel synergistic and highly effective drug combination for the treatment of castration-resistant prostate cancer.

## Discussion

The clinical significance of the non-protein coding genome in prostate cancer is rapidly gaining attention. Recently, whole-genome sequencing of primary prostate cancer specimens revealed enrichment of somatic mutations in AR chromatin binding sites^59,60^, a subset of which functionally affected enhancer activity^15^. Not only in primary prostate cancer but also in the mCRPC setting, non-coding somatic alterations have been reported, including amplification of enhancer elements that regulate expression of AR^61,62^, HOXB13^18^ and FOXA1^18^. In addition to somatic alterations in the primary DNA sequence or copy number, modifications in epigenetic regulatory elements are proving crucial in prostate cancer development and progression. Furthermore, extensive epigenetic reprogramming and AR enhancer plasticity have been related to tumorigenesis^15,16^ and progression^18^, as well as therapy resistance^17^. To date, deviations in enhancer regulation have not been extensively studied in castration-resistant disease and have not been explored in the context of a controlled clinical trial. Here, we interrogated the epigenome in relation to AR-targeted therapy response in mCRPC patients. Within our clinical cohort, epigenetic features revealed a robust classification scheme predicting response to treatment. These findings additionally revealed potential drivers of resistance as well as novel therapeutic drug combinations to combat castration-resistant disease.

While ENZA improves outcome in patients with mCRPC^12,13^, a significant proportion of mCRPC patients experience no response to AR-targeting treatment due to intrinsic resistance mechanisms. Supporting the notion of a pre-existing treatment resistance patient population, we found that H3K27ac profiles in mCRPC tumors remained unaltered following ENZA treatment. These data suggest that these cancers were already resistant prior to drug exposure, harboring epigenetic programs that support AR-independent cellular growth. In contrast, most previously described resistance mechanisms appeared to be treatment-induced, including AR mutations^63-65^ and amplification^40^, GR upregulation^47-49^ or enrichment of HNF4G^46^. As the tumor samples we analyzed were already relapsed after prior therapies and developed castration resistance, it is plausible that the induction of the abovementioned resistance mechanisms occurred before our samples were taken. At the single-cell level, genes associated with our H3K27ac non-responder sites were significantly enriched in an ENZA-resistance-associated cluster which expands after long-term treatment^32^, indicating that the acetylated regions we have identified – and associated genes – are important in driving therapeutic resistance in a subset of tumors. These results may jointly point towards the induction of divergent castration resistance mechanisms, which either sustain hormone-dependency (as in the case for our ‘responder’ population) or diverge towards complete hormone-independence (our ‘non-responder’ population), in which other transcription factors compensate for the lack of AR activity.

Apart from the hormone receptor family, transcription factors are generally considered challenging drug targets and potential other therapeutic strategies – such as epigenetic drugs – may prove of clinical benefit for these cases. Along these lines, for three of our hits: NKX3-1, FOXA1 and GATA2, specific inhibitors are yet to be developed. Recently, indirect small molecule inhibition of FOXA1 has been described, by means of targeting EZH2^66^ and LSD1^67^, presenting a potential direct therapeutic avenue in this setting.

As HDAC3 expression has been shown to be critical for AR-driven transcriptional programs – both in hormone-sensitive and castration-resistant cell line models^68^ – we chose to further study the potential benefit of HDAC inhibition as a therapeutic strategy in mCRPC. Our data reveal that HDAC3 is also critically involved in resistance to AR-targeted therapeutics in the mCRPC setting, and prove therapeutic proof-of-concept of a synergistic drug-drug interaction between ENZA and the pan-HDAC inhibitor vorinostat, both in cell line models and mCRPC PDX-derived explants. This drug has been clinically approved in the treatment of cutaneous T cell lymphomas and also shown promise as a therapeutic strategy in advanced non-small cell lung cancer^69^. Although, vorinostat showed no activity as a single agent in mCRPC patients, a phase 2 trial into the combination of the non-selective HDAC inhibitor panobinostat and the AR targeted drug bicalutamide in 55 patients, showed promising results^70,71^. Common drug-related serious adverse events such as thromboembolic events associated with vorinostat are of concern^55^, but may be avoided with the likely lower concentrations used in a combination with ENZA.

## Conclusions

Based on our results, new clinical trials for testing vorinostat – or other HDAC inhibitors – in conjunction with ENZA for mCRPC patients would be justified, since novel highly-effective drug-drug combinations are urgently needed to combat this deadly disease.

## Data Availability

Raw ChIP-sequence data are deposited in the European Genome-Phenome Archive (EGAS00001006161).

https://ega-archive.org/studies/EGAS00001006161

## Declarations

### Ethical Approval and Consent to Participate

The trial was approved by the institutional review board of the Netherlands Cancer Institute, written informed consent was signed by all participants enrolled in the study, and all research was carried out in accordance with relevant guidelines and (inter-)national and ethical standards. Within the General Data Protection Regulation, patients always had the opportunity to object or actively consent to the (continued) use of their personal data and biospecimens for research purposes.

### Consent for Publication

All authors have read the manuscript and consent to publication.

### Availability of Supporting Data

Raw ChIP-sequence data are deposited in the European Genome-Phenome Archive (EGAS00001006161)

### Authors’ Contributions

TMS, YZ, SP, WZ and AMB wrote the main manuscript text. TMS, YZ, SP, KS, HMN, LGB, SH, YK, JK, SL, SS, CL, VvdN, JS, BM, GJ, GFJLvL, AU, RLB, EC, WZ and AMB prepared and analyzed data for all figures and tables. All other authors reviewed the manuscript.

## Acknowledgments

The authors gratefully acknowledge the patients and the families of patients who contributed to this study. We thank the NKI Genomics Core Facility, Core Facility Molecular Pathology and Biobanking, Research High Performance Computing Facility, Scientific Information Service for their excellent technical support and all group members from the Zwart, Bergman and Corey labs for highly constructive feedback and suggestions. The authors thank the A.U. Norwegian Cancer Society, Academy of Finland and Cancer Foundation Finland, S.H. and

M.N. Academy of Finland and Sigrid Jusélius Foundation, Finnish Cancer Institute.

## Supplemental information

**Supplemental File 1**

Contains 9 Supplementary Figures and 4 Supplementary Tables

**Supplemental File 2**

Contains additional clinical trial information

