## Supplementary Figure for "Enhancer profiling identifies epigenetic markers of endocrine resistance and reveals therapeutic options for metastatic castration-resistant prostate cancer patients"

The supplementary file includes 9 Figures and 5 Tables.

Tesa M. Severson[1,2,3,*], Yanyun Zhu[1,2,*], Stefan Prekovic[1,2,*], Karianne Schuurman[1], Holly M. Nguyen[4], Lisha G. Brown[4], Sini Hakkola[5], Yongsoo Kim[1,6], Jeroen Kneppers[1,2], Simon Linder[1,2], Suzan Stelloo[1,2,7], Cor Lieftink[3], Michiel van der Heijden[3,8], Matti Nykter[5], Vincent van der Noort[9], Joyce Sanders[10], Ben Morris[3], Guido Jenster[11], Geert JLH van Leenders[12], Mark Pomerantz[13], Matthew L. Freedman[13,14], Roderick L. Beijersbergen[6], Alfonso Urbanucci[5,15], Lodewyk Wessels[2,3,16], Eva Corey[4], Wilbert Zwart[1,2,17#], Andries M. Bergman[1,8#]

**1** Division of Oncogenomics, The Netherlands Cancer Institute, Amsterdam, the Netherlands
**2** Oncode Institute, the Netherlands
**3** Division of Molecular Carcinogenesis, The Netherlands Cancer Institute, Amsterdam, the Netherlands
**4** Department of Urology, University of Washington, Seattle, WA, USA
**5** Prostate Cancer Research Center, Faculty of Medicine and Health Technology, Tampere University and Tays Cancer Centre, Tampere, Finland
**6** present working address: Department of Pathology, Cancer Center Amsterdam, Amsterdam UMC, Vrije Universiteit Amsterdam, Amsterdam, the Netherlands
**7** present working address: Department of Molecular Biology, Faculty of Science, Radboud Institute for Molecular Life Sciences, Oncode Institute, Radboud University Nijmegen, 6525 GA Nijmegen, The Netherlands.
**8** Division of Medical Oncology, The Netherlands Cancer Institute, Amsterdam, the Netherlands
**9** Department of Biometrics, The Netherlands Cancer Institute, Amsterdam, The Netherlands.
**10** Department of Pathology, The Netherlands Cancer Institute, Amsterdam, the Netherlands
**11** Department of Urology, Erasmus MC, Rotterdam, The Netherlands
**12** Department of Pathology, Erasmus MC Cancer Institute, University Medical Centre, Rotterdam, The Netherlands.
**13** Department of Medical Oncology, Dana-Farber Cancer Institute, Harvard Medical School, Boston, MA, USA.
**14** The Eli and Edythe L. Broad Institute, Cambridge, MA, USA
**15** Department of Tumor Biology, Institute for Cancer Research, Oslo University Hospital, Oslo, Norway
**16** Department of EEMCS, Delft University of Technology, Delft, The Netherlands.
**17** Laboratory of Chemical Biology and Institute for Complex Molecular Systems, Department of Biomedical Engineering, Eindhoven University of Technology, Eindhoven, The Netherlands.

*shared first authors

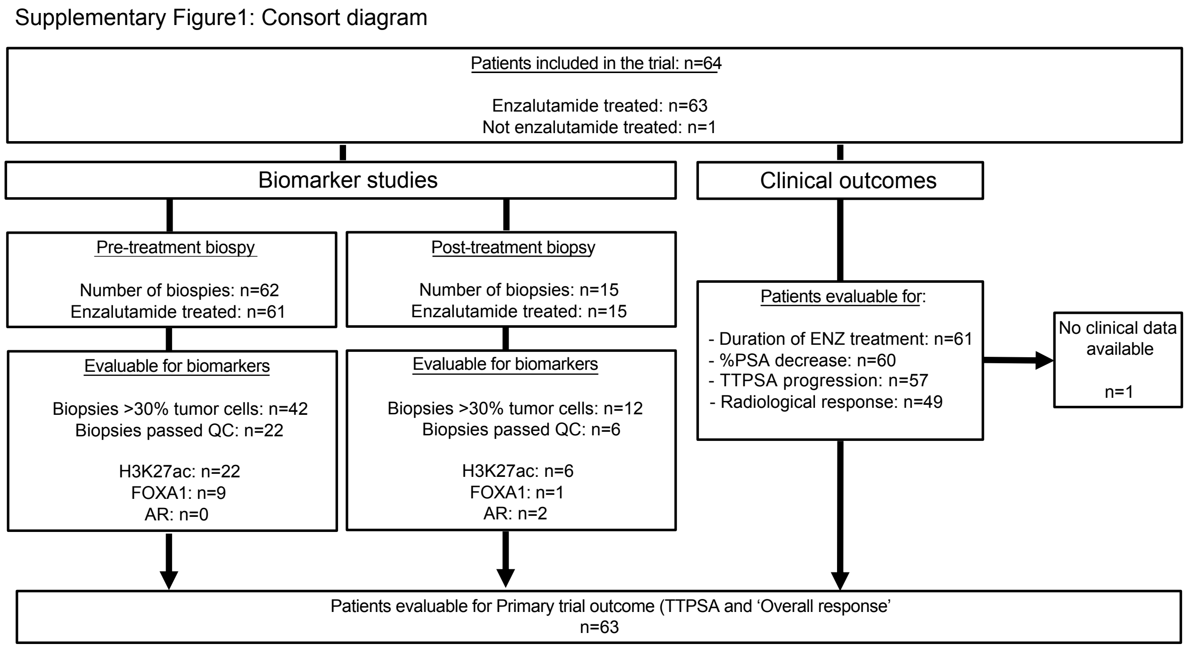

**Supplementary Fig. 1** CONSORT diagram displaying the patient and biomarker information for the trial

Supplementary Figure 2: Quality control metrics for H3K27ac ChIP-seq data from mCRPC samples

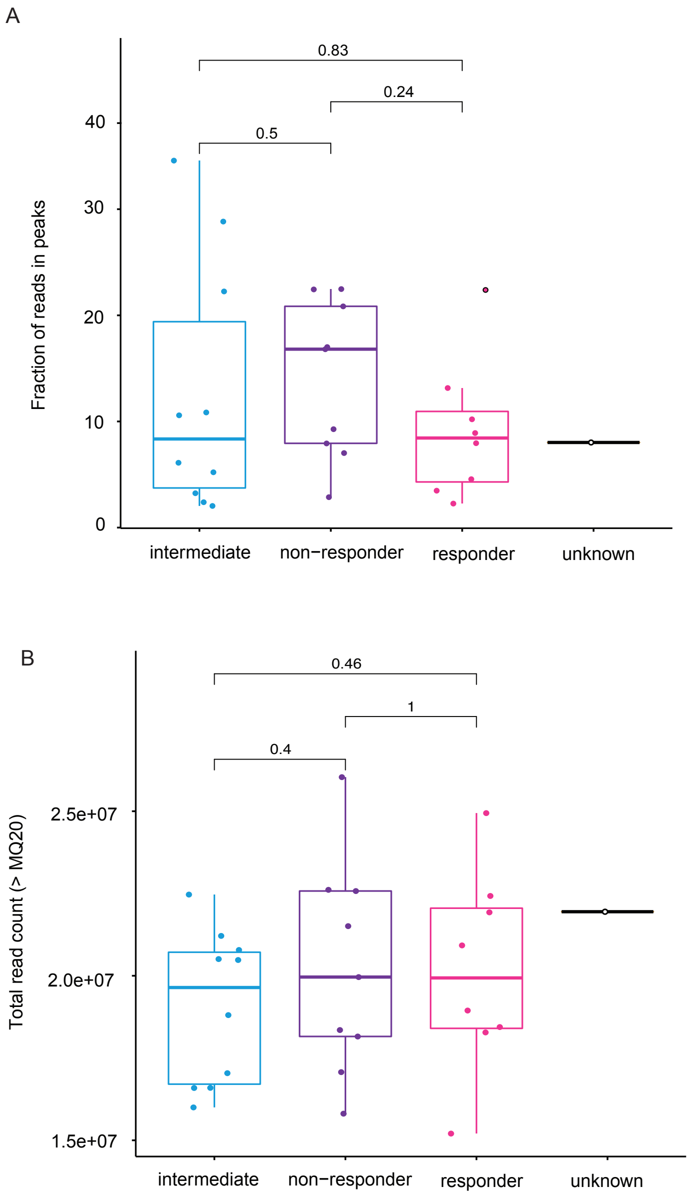

**Supplementary Fig. 2** Quality control metrics for H3K27ac ChIP-seq data from mCRPC samples. **(A)** Boxplots of the fraction of reads in peaks (FRiP) for H3K27ac ChIP-seq samples (n=28). Sample type depicted on the x-axis, FRiP on y-axis. Outlier point is indicated with black outline. For both panels, boxes indicate interquartile range (IQR, 25-75%) of the data. Box center line is the median. Whiskers extend to the minimum or maximum value no more than 1.5 time the IQR from the top or bottom of the box, respectively. **(B)** Boxplots of the total read count (>MQ20) for H3K27ac ChIP-seq samples (n=28). Sample type depicted on the x-axis, read count on y-axis.

Supplementary Figure 3: PCA of H3K27ac profiles with sample type labels

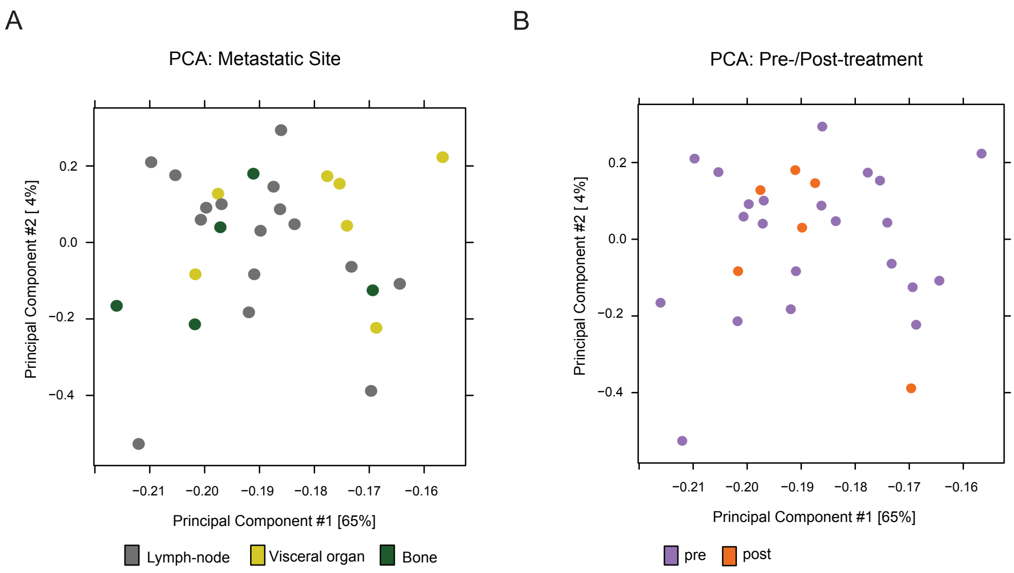

**Supplementary Fig. 3** Principal Component Analysis (PCA) for H3K27ac profiles with sample type labels **(A)** PCA using normalized read counts in all peaks in H3K27ac ChIP-seq data (n=73039) for all samples (n=28) with metastatic site labeled: lymph-node (grey), visceral organ (yellow) and bone (black). **(B)** PCA using normalized read counts in all peaks in H3K27ac ChIP-seq data (n=73039) for all samples (n=28) with treatment status labeled: pre-treatment (purple) or post-treatment (orange).

Supplementary Figure 4: H3K27ac ChIP-seq data analyses in primary prostate cancers, metastatic prostate cancer and normal tissue focused on the 657 mCRPC resistance-associated regions

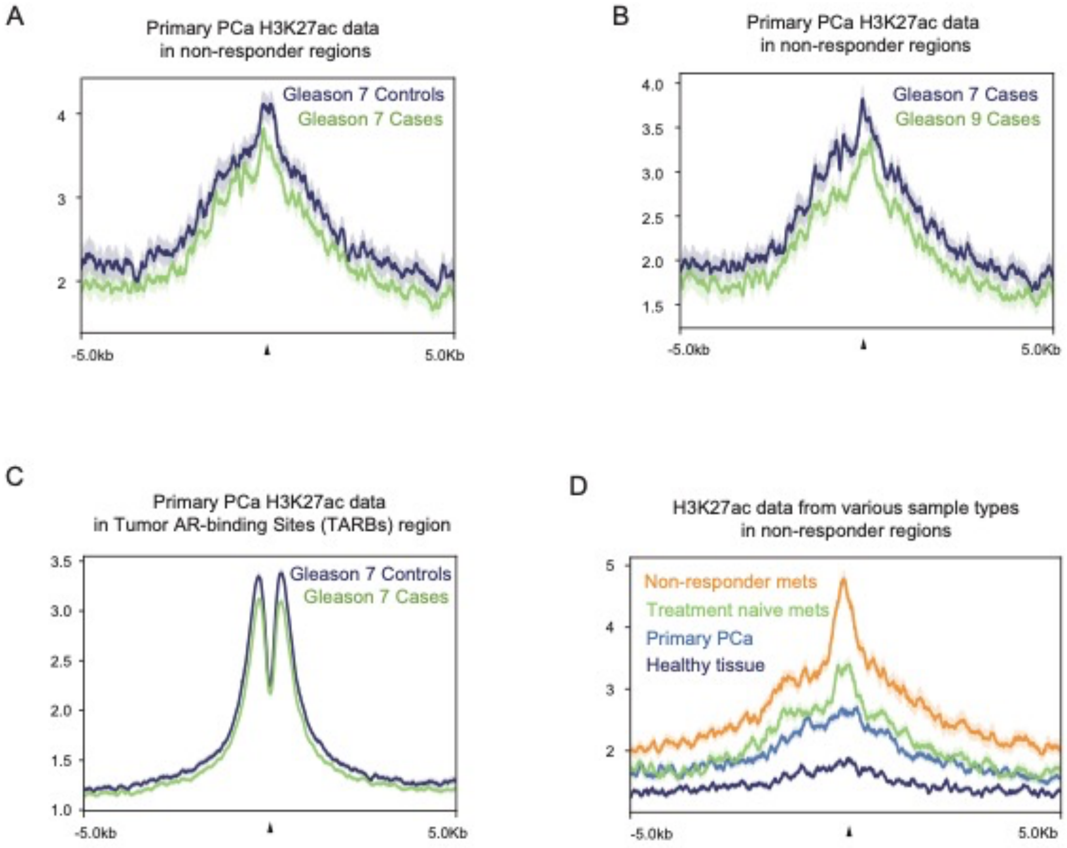

**Supplementary Fig. 4** H3K27ac ChIP-seq data analyses in primary prostate cancer, metastatic prostate cancer and normal tissue focused on the 657 mCRPC resistance-associated regions. **(A)** Average H3K27ac read count profiles of merged data at the 657 mCRPC resistance-associated H3K27ac sites (±5 kb from the peak center), comparing Gleason 7 primary tumors that progressed within 5 years (Cases, green, n=24) or remained free from progression for at least 15 years (Controls, blue, n=20). **(B)** Average H3K27ac read count profiles of merged data at the 657 mCRPC resistance-associated H3K27ac sites (±5 kb from the peak center), comparing primary tumor data from Cases which progressed within 5 years and were either Gleason 7 (blue, n=24) or Gleason 9 (green, n=12). **(C)** Average H3K27ac read count profiles of merged data at Tumor AR binding sites (TARBS, n=9182) (±5 kb from the peak center), comparing Gleason 7 primary tumors that progressed within 5 years (Cases, green, n=24) or remained free from progression for at least 15 years (Controls, blue, n=20). **(D)** Average H3K27ac read count profiles of merged data at the 657 mCRPC resistance-associated H3K27ac sites (±5 kb from the peak center), comparing metastatic non-responder data (this study, orange, n=9), treatment-naïve metastatic data (green, n=19), data from primary prostate (n=9) and data from healthy prostate tissue (n=9). For all panels, shading indicates standard-error of the data.

Supplementary Figure 5: H3K27ac ChIP-seq global correlation among PDX samples

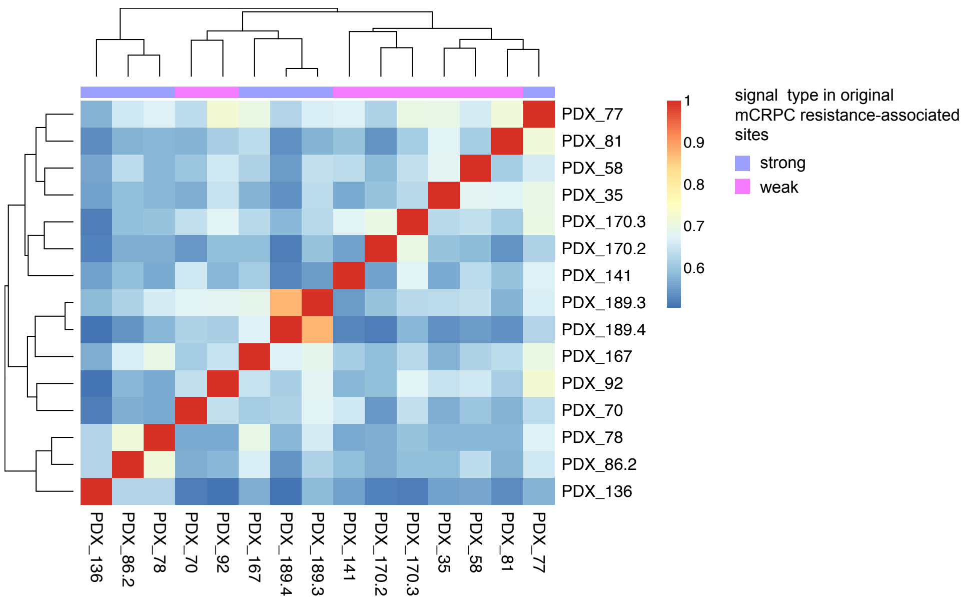

**Supplementary Fig. 5** H3K27ac ChIP-seq global correlation among PDX samples. Correlation of H3K27ac ChIP-seq signal (50kb bins across the genome, Pearson correlation) in mCRPC PDX samples (n=15). Indicated is class of samples based on signal intensity at the 657 H3K27ac mCRPC resistance-associated regions: weak (pink) or strong (purple).

Supplementary Figure 6: *In vivo* response to castration in mCRPC PDX models

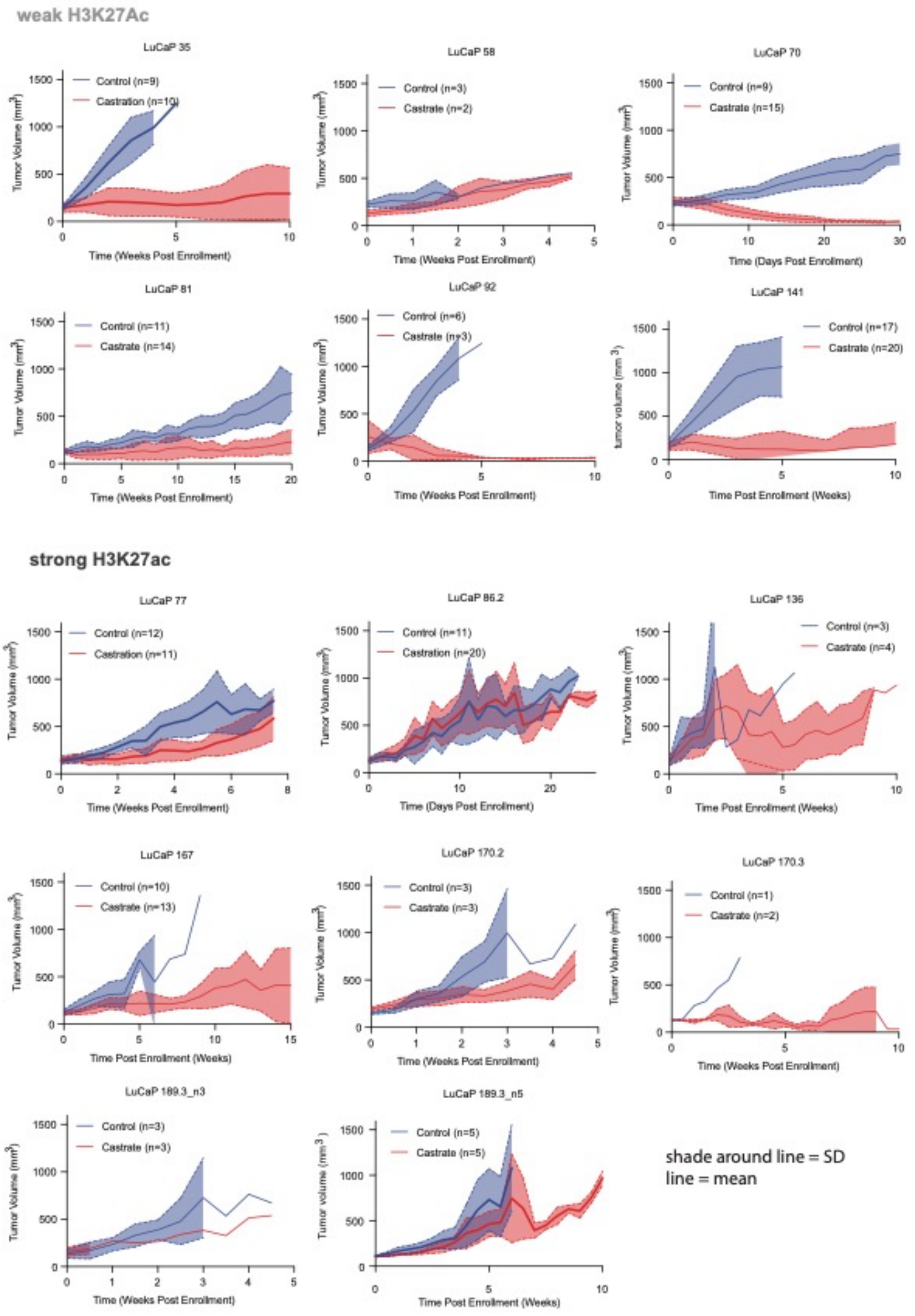

**Supplementary Fig. 6** *In vivo* response to castration in mCRPC PDX models (Top) and (bottom) groups indicate those models with weak or strong H3K27ac signal in 657 mCRPC resistance-associated regions, respectively. For each model, tumor volume (in $mm^3$) (y-axis) is indicated in time, for control (blue) or castrated (red) conditions. Time is shown on the x-axis. Solid line indicates the mean of the tumor volume. Shaded area around the solid line indicates the standard deviation of the mean (solid line). Number of animals per group is shown per panel

Supplementary Figure 7: Knockdown confirmation of siRNA deconvolution experiments

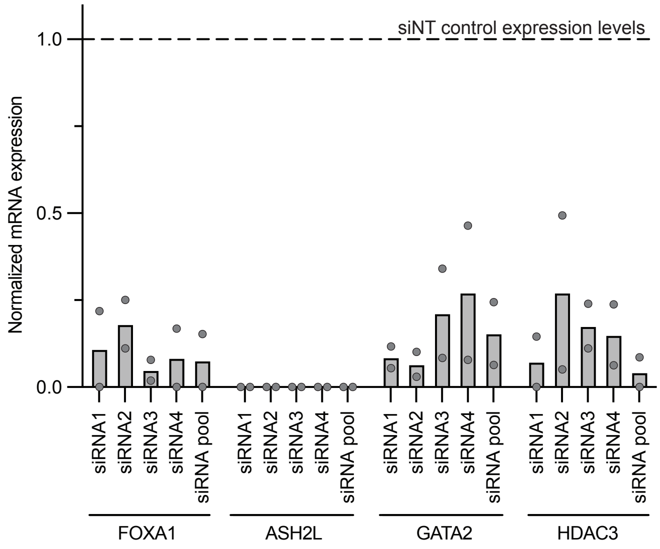

**Supplementary Fig. 7** Knockdown confirmation of siRNA deconvolution experiments. Expression levels are shown for each siRNA separately, from 2 biological replicates with 2 technical replicates each. Data are normalized over siControl. Dots indicate the mean of each technical replicate. The bar indicates the mean of these values.

Supplementary Figure 8: Sensitivity to HDAC inhibition

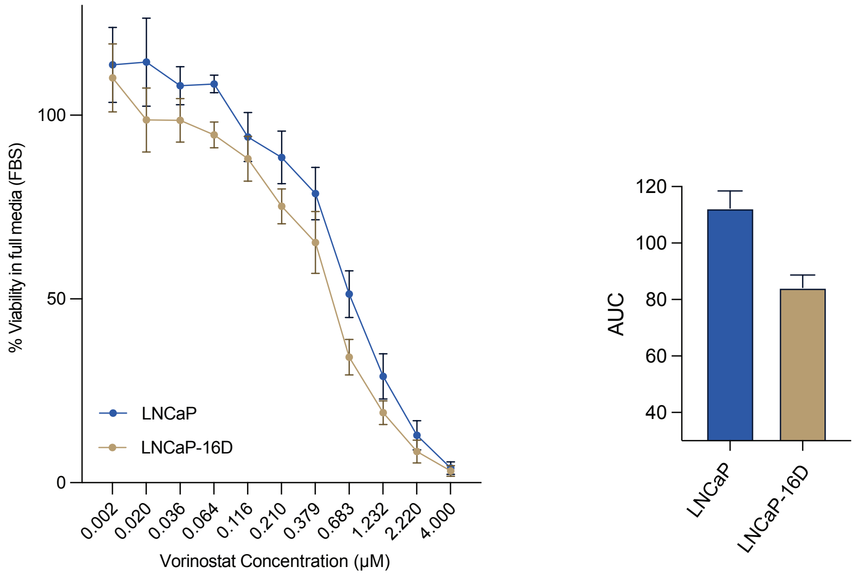

**Supplementary Fig. 8** Sensitivity to HDAC inhibition. (Left) Sensitivity of LNCaP and LNCaP-16D cells to vorinostat alone. LNCaP (blue) and LNCaP-16D (gold) cells are exposed to increasing concentrations of vorinostat, and cell viability is determined in full medium (FBS). Error bars indicated SEM from 4 biological replicates. (Right) Area under the curve (AUC) for both cell lines. Error bars indicate SD from 4 biological replicates.

Supplementary Figure 9: Drug sensitivity and synergy analyses for cell-lines and mCRPC PDX explants

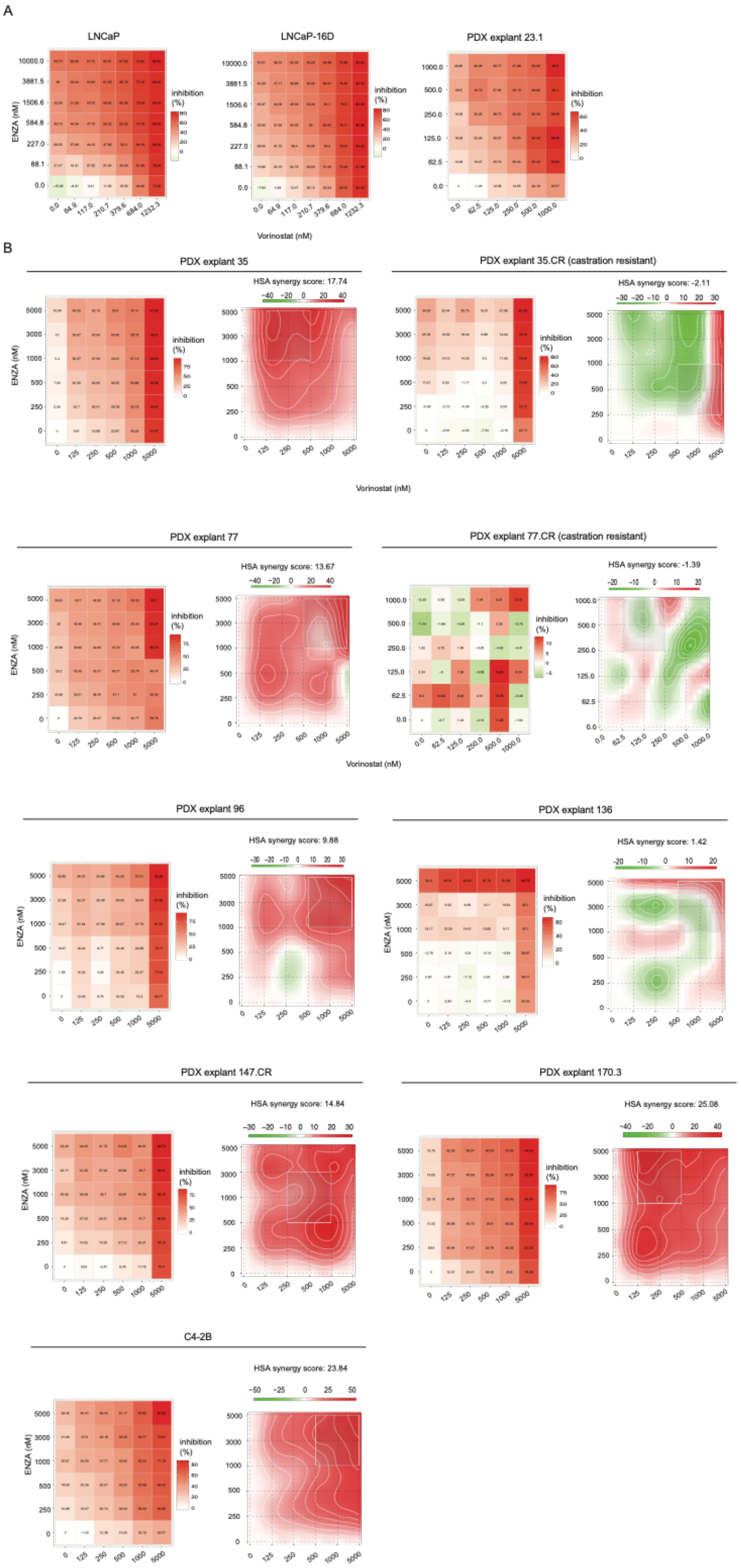

**Supplementary Fig. 9** Drug sensitivity and synergy analyses for cell-lines and mCRPC PDX explants. **(A)** Drug sensitivity data for synergy data shown in Figure 4. For all cell-lines and mCRPC PDX models, explants were generated and exposed to increasing concentrations of vorinostat (x-axis) or enzalutamide (ENZA) (y-axis) and assessed for viability. Percent inhibition of growth is shown from low (green) to high (red). **(B)** (Left) Drug sensitivity for additional mCRPC models and cell-line (C4-2B) as above. (Right) Drug synergy representation for mCRPC PDX explants and cell-line. Synergy determined using the HAS model (score > 10 indicates synergy with regions of maximal synergy outlined in white).

#### Supplementary Table 1. Baseline Characteristics of mCRPC patients with a pre-enzalutamide treatment biopsy

*p:

| Patient demographics | Median or value [IQR], Number of Patients (%) | | | | | | | |
|---|---|---|---|---|---|---|---|---|
| | | | Pre-treament biopsies | | | Post-treatment biopsies | | |
| | Whole population (n=64) | | Evaluable population (n=22) | | P* | Evaluable Population (n=6) | | P** |
| Age (years) | | | | | | | | |
| | 69 | [64-73] | 68 | [63-72] | 0.71 | 66 | [63-72] | 0.71 |
| ECOG performance status (number of patients) | | | | | | | | |
| 0 | 10 | 15.6% | 6 | 27.3% | | 0 | 0.0% | |
| 1-2 | 54 | 84.4% | 16 | 77.7% | | 6 | 100.0% | |
| Gleason score (number of patients) | | | | | | | | |
| 6 | 5 | 7.8% | 2 | 9.1% | | 0 | 0.0% | |
| 7 | 19 | 29.7% | 10 | 45.5% | | 3 | 50.0% | |
| 8-10 | 37 | 57.8% | 9 | 40.9% | | 3 | 50.0% | |
| Missing data | 3 | 4.7% | 1 | 4.5% | | 0 | 0.0% | |
| Metastatic sites (number of patients) | | | | | | | | |
| Bone | 48 | 75.0% | 15 | 68.2% | | 5 | 83.3% | |
| Lymph nodes | 45 | 70.3% | 17 | 77.3% | | 4 | 66.7% | |
| Visceral organs | 22 | 34.4% | 7 | 31.8% | | 2 | 33.3% | |
| PSA prior to treatment (µg/l) | | | | | | | | |
| | 55.9 | [86-148] | 94.5 | [47-262] | 0.81 | 21.3 | [9-46] | 0.45 |
| PSA progression prior to treatment | | | | | | | | |
| Yes | 60 | 93.8% | 21 | 95.5% | | 6 | 100.0% | |
| No | 3 | 4.7% | | 4.5% | | 0 | 0.0% | |
| Not evaluable | 1 | 1.6% | 0 | 0.0% | | 0 | 0.0% | |
| Radiological progression prior to treatment | | | | | | | | |
| Yes | 52 | 81.3% | 20 | 90.9% | | 5 | 83.3% | |
| No | 11 | 17.2% | 2 | 9.1% | | 1 | 16.7% | |
| Not evaluable | 1 | 1.6% | 0 | 0.0% | | 0 | 0.0% | |
| Biopsy side | | | | | | | | |
| Bone | 19 | 29.7% | 4 | 18.2% | | 1 | 16.7% | |
| Lymph nodes | 32 | 50.0% | 13 | 59.1% | | 3 | 50.0% | |
| Visceral organs | 13 | 20.3% | 5 | 22.7% | | 2 | 33.3% | |
| Line of mCRPC treatment | | | | | | | | |
| 1 | 21 | 32.8% | 6 | 27.3% | | 1 | 16.7% | |
| 2 | 19 | 29.7% | 6 | 27.3% | | 4 | 66.7% | |
| >2 | 24 | 37.5% | 10 | 45.4% | | 1 | 16.7% | |
| Prior treatments | | | | | | | | |
| Taxane drugs | 40 | 62.5% | 15 | 68.2% | | 5 | 83.3% | |
| ARTA | 15 | 23.4% | 10 | 45.5% | | 1 | 16.7% | |

comparison pre-treatment evaluable population with whole population (Student T-test)

**p: comparison post-treatment evaluable population with whole population (Student T-test)
mCRPC: metastatic Castration Resistant Prostate Cancer
IQR: Inter Quartile Range
ECOG: Eastern Cooperative Oncology Group
PSA: Prostate Specific Antigen
Taxane drugs: docetaxel and/or cabazitaxel
ARTA: Androgen Receptor Targeting Agent

**Supplementary Table 2. Outcomes of enzalutamide treatment of mCRPC patients with a pre-enzalutamide treatment biopsy**

| Patient demographics | Median or value [IQR], Number of Patients (%) | | | | | | | |
|---|---|---|---|---|---|---|---|---|
| | | | Pre-treatment biopsies | | | Post-treatment biopsies | | |
| | Whole population (n=64) | | Evaluable population (n=22) | | P* | Evaluable population (n=6) | | P** |
| Follow-up (months) | 17 | [9-30] | 12 | [6-20] | 0.14 | 25 | [17-33] | 0.29 |
| Enzalutamide treated (number of patients) | | | | | | | | |
| Yes | 63 | 98.4% | 21 | 95.5% | | 6 | 100.0% | |
| No | 1 | 1.6% | 1 | 4.5% | | 0 | 0.0% | |
| Enzalutamide dose (mg/day) | 160 | [160-160] | 160 | [160-160] | 0.28 | 160 | [160-160] | 0.68 |
| Duration of treatment (weeks) | 22.7 | [13-45] | 22.9 | [14-40] | 0.95 | 34.6 | [16-90] | 0.19 |
| PSA change from baseline (%) | 45.5 | [12-73] | 48.2 | [16-77] | 0.73 | 50.2 | [43-66] | 0.84 |
| 50% PSA decrease from baseline | | | | | | | | |
| Yes | 30 | 46.9% | 10 | 45.5% | | 3 | 50.0% | |
| No | 29 | 45.3% | 10 | 45.5% | | 2 | 33.3% | |
| Not evaluable | 5 | 7.8% | 2 | 9.1% | | 1 | 16.7% | |
| Time to PSA progression (weeks) | 17.7 | [12-34] | 16.4 | [11-35] | 0.78 | 24.6 | [14-27] | 0.82 |
| Not evaluable | 7 | 10.9% | 2 | 9.1% | | 1 | 16.7% | |
| Radiological response | | | | | | | | |
| CR/PR | 12 | 18.8% | 8 | 36.4% | | 0 | 0.0% | |
| SD | 11 | 17.2% | 0 | 0.0% | | 3 | 50.0% | |
| PD | 26 | 40.6% | 11 | 50.0% | | 2 | 33.3% | |
| Not evaluable | 15 | 23.4% | 3 | 13.6% | | 1 | 16.7% | |
| Overall response | | | | | | | | |
| Response | 15 | 23.4% | 6 | 27.3% | | 2 | 33.3% | |
| Intermediate | 24 | 37.5% | 8 | 36.4% | | 2 | 33.3% | |
| No response | 24 | 37.5% | 8 | 36.4% | | 1 | 16.7% | |
| Not evaluable | 1 | 1.6% | 0 | 0.0% | | 1 | 16.7% | |
| Overall survival (months) | 14.5 | [8-26] | 10 | [5-15] | 0.19 | 24.5 | [17-33] | 0.20 |
| Patients alive at the close of database | 8 | 12.5% | 3 | 13.6% | | 0 | 0.0% | |

*p: comaparison pre-treatment evaluable population with whole population (Student T-test)

**p: comaparison post-treatment evaluable population with whole population (Student T-test)
mCRPC: metastatic Castration Resistant Prostate Cancer
IQR: Inter Quartile Range
PSA: Prostate Specific Antigen
CR: Complete Response, PR: Partial Response, SD: Stable Disease, PD: Progressive Disease

**Supplementary Table 3. Number of peaks (MACS2), RSC (relative strand cross-correlation coefficient) values, read counts and Fraction of Reads in Peaks (FRiP) values for all ChIP-seq datasets**

| NKI_ID | number_peaks | RSC_value | FRiP | total_MQ20_readcount | factor | response | EGA_EGAS00001006161_alias_ID |
|---|---|---|---|---|---|---|---|
| wz3535 | 38569 | 1.31163 | 9.08 | 21607078 | AR | intermediate | patient35_post_AR |
| wz3537 | 8568 | 0.83715 | 1.57 | 15847289 | AR | unknown | patient68_post_AR |
| wz3018 | 11840 | 1.04127 | 1.76 | 27703555 | FOXA1 | non-response | patient42_pre_FOXA1 |
| wz3019 | 18127 | 1.15026 | 3.81 | 21857548 | FOXA1 | non-response | patient41_pre_FOXA1 |
| wz3184 | 21123 | 0.95601 | 4.52 | 11443399 | FOXA1 | intermediate | patient35_pre_FOXA1 |
| wz3185 | 19197 | 1.24551 | 3.96 | 21610894 | FOXA1 | response | patient48_pre_FOXA1 |
| wz3203 | 11316 | 1.02411 | 2.9 | 13845140 | FOXA1 | response | patient49_pre_FOXA1 |
| wz3205 | 34042 | 1.28796 | 7.61 | 21404943 | FOXA1 | non-response | patient54_pre_FOXA1 |
| wz3211 | 10468 | 0.97813 | 2.45 | 15756501 | FOXA1 | intermediate | patient37_pre_FOXA1 |
| wz3226 | 19226 | 1.22192 | 4.71 | 22055352 | FOXA1 | non-response | patient60_pre_FOXA1 |
| wz3539 | 14255 | 0.87093 | 2.45 | 19331353 | FOXA1 | intermediate | patient35_post_FOXA1 |
| wz3541 | 11859 | 1.14908 | 3.26 | 24328956 | FOXA1 | response | patient67_pre_FOXA1 |
| wz2925 | 50143 | 0.96048 | 10.85 | 21212042 | H3K27ac | intermediate | patient9_pre_H3K27ac |
| wz2926 | 13529 | 0.89983 | 2.04 | 17040576 | H3K27ac | intermediate | patient17_pre_H3K27ac |
| wz3024 | 64991 | 1.10667 | 17.01 | 26036673 | H3K27ac | non-response | patient42_pre_H3K27ac |
| wz3025 | 59168 | 1.2257 | 20.85 | 18155445 | H3K27ac | non-response | patient41_pre_H3K27ac |
| wz3026 | 47457 | 1.06629 | 10.21 | 24944506 | H3K27ac | response | patient20_post_H3K27ac |
| wz3130 | 22078 | 1.10244 | 6.11 | 16000146 | H3K27ac | intermediate | patient12_pre_H3K27ac |
| wz3131 | 12889 | 0.98111 | 3.49 | 15206340 | H3K27ac | response | patient26_pre_H3K27ac |
| wz3167 | 29791 | 1.13127 | 7.94 | 17074034 | H3K27ac | non-response | patient23_pre_H3K27ac |
| wz3173 | 13325 | 0.72371 | 2.87 | 15810807 | H3K27ac | non-response | patient11_pre_H3K27ac |
| wz3187 | 100760 | 1.30923 | 28.84 | 20480022 | H3K27ac | intermediate | patient35_pre_H3K27ac |
| wz3189 | 36040 | 1.02221 | 7.96 | 22427023 | H3K27ac | response | patient48_pre_H3K27ac |
| wz3190 | 44749 | 1.00845 | 8.92 | 21929446 | H3K27ac | response | patient56_pre_H3K27ac |
| wz3200 | 35744 | 0.93106 | 7.03 | 19961048 | H3K27ac | non-response | patient25_post_H3K27ac |
| wz3206 | 73022 | 1.31309 | 22.39 | 18442118 | H3K27ac | response | patient49_pre_H3K27ac |
| wz3207 | 44026 | 1.12156 | 9.28 | 21509920 | H3K27ac | non-response | patient53_pre_H3K27ac |
| wz3208 | 68492 | 1.30751 | 22.48 | 22575845 | H3K27ac | non-response | patient54_pre_H3K27ac |
| wz3214 | 82775 | 1.33305 | 22.25 | 20783989 | H3K27ac | intermediate | patient37_pre_H3K27ac |
| wz3220 | 63902 | 1.13743 | 13.15 | 20922983 | H3K27ac | response | patient63_post_H3K27ac |
| wz3227 | 23086 | 0.90727 | 5.21 | 18804886 | H3K27ac | intermediate | patient13_pre_H3K27ac |
| wz3228 | 12823 | 0.76617 | 2.4 | 20509302 | H3K27ac | intermediate | patient29_pre_H3K27ac |
| wz3230 | 54349 | 1.29656 | 22.44 | 22612193 | H3K27ac | non-response | patient60_pre_H3K27ac |
| wz3396 | 10710 | 0.97102 | 3.24 | 16594416 | H3K27ac | intermediate | patient16_post_H3K27ac |
| wz3397 | 11809 | 0.96084 | 2.27 | 18945269 | H3K27ac | response | patient52_pre_H3K27ac |
| wz3542 | 65764 | 1.41907 | 34.59 | 22469696 | H3K27ac | intermediate | patient35_post_H3K27ac |
| wz3543 | 40453 | 1.079 | 16.81 | 18352062 | H3K27ac | non-response | patient65_pre_H3K27ac |
| wz3544 | 35446 | 1.10497 | 8.02 | 21946402 | H3K27ac | unknown | patient68_post_H3K27ac |
| wz3545 | 15551 | 0.99225 | 4.57 | 18280001 | H3K27ac | response | patient67_pre_H3K27ac |
| wz678 | 48391 | 1.01082 | 10.59 | 16591501 | H3K27ac | intermediate | patient7_pre_H3K27ac |

**Supplementary Table 4. Genomic features annotation of differential peaksets.**

| Annotated peaks generated by ChIPsee | | |
|---|---|---|
| # Non-responder enriched peaks | | |
| # 657/657 peaks were annotated | | |
| # Genomic Annotation Summary: | | |
| Feature | Frequency | |
| Promoter | 3.805175 | |
| 5' UTR | 0.608828 | |
| 3' UTR | 1.826484 | |
| 1st Exon | 0.608828 | |
| Other Exon | 5.936073 | |
| 1st Intron | 12.024353 | |
| Other Intron | 36.225266 | |
| Downstream | 0.761035 | |
| Distal Interge | 38.203957 | |
| # Responder_enriched | | |
| # 25/25 peaks were annotated | | |
| # Genomic Annotation Summary: | | |
| Feature | Frequency | |
| Promoter | | 4 |
| Other Exon | | 8 |
| 1st Intron | | 24 |
| Other Intron | | 40 |
| Downstream (<=300) | | 4 |
| Distal Intergenic | | 20 |

**Supplementary Table 5. List of Public ChIP-seq data sets queried for GIGGLE enrichment analysis and results**

| CistromeDB human prostate specific experiments | | | | Giggle results | | | |
|---|---|---|---|---|---|---|---|
| GEO_ID | Cell_type | Factor | | Factor | median_GIGGLE_combo_score | rank | siRNA_focused_screen |
| GSM353640 | 22RV1 | AR | | ZMYND8 | -0.081722 | 1 | FALSE |
| GSM759660 | LNCaP | AR | | EED | -0.008737 | 2 | FALSE |
| GSM759659 | LNCaP | AR | | EP300 | 0 | 3 | FALSE |
| GSM759658 | LNCaP | AR | | MED12 | 0 | 4 | FALSE |
| GSM759657 | LNCaP | AR | | SUZ12 | 0 | 5 | FALSE |
| GSM696839 | LNCaP | AR | | E2F1 | 0 | 6 | FALSE |
| GSM353644 | LNCaP | AR | | KDM1A | 0 | 7 | FALSE |
| GSM353642 | LNCaP | AR | | CTBP1 | 0 | 8 | FALSE |
| GSM353641 | LNCaP | AR | | CTBP2 | 0 | 9 | FALSE |
| GSM686944 | LNCaP | EP300 | | FOXP1 | 0 | 10 | FALSE |
| GSM686943 | LNCaP | EP300 | | CHD1 | 0 | 11 | FALSE |
| GSM353648 | LNCaP | ERG | | SMARCA4 | 0 | 12 | FALSE |
| GSM759664 | LNCaP | FOXA1 | | NANOG | 0 | 13 | FALSE |
| GSM759663 | LNCaP | FOXA1 | | CCNT1 | 0 | 14 | FALSE |
| GSM759662 | LNCaP | FOXA1 | | WDHD1 | 0 | 15 | FALSE |
| GSM759661 | LNCaP | FOXA1 | | ZFX | 0 | 16 | FALSE |
| GSM353633 | LNCaP | FOXA1 | | TRIM28 | 0 | 17 | FALSE |
| GSM686942 | LNCaP | H2AZ | | CTCF | 0.031704 | 18 | FALSE |
| GSM686941 | LNCaP | H2AZ | | BMI1 | 0.088678 | 19 | FALSE |
| GSM686946 | LNCaP | MED12 | | POU2F1 | 0.13813 | 20 | FALSE |
| GSM686945 | LNCaP | MED12 | | ETS1 | 0.233216 | 21 | FALSE |
| GSM759670 | LNCaP | NR3C1 | | JUND | 0.263972 | 22 | FALSE |
| GSM759669 | LNCaP | NR3C1 | | WDR5 | 0.554446 | 23 | FALSE |
| GSM696844 | LNCaP | POLR2A | | SFPQ | 0.572018 | 24 | FALSE |
| GSM696843 | LNCaP | POLR2A | | DNMT1 | 0.68868 | 25 | FALSE |
| GSM353618 | LNCaP | POLR2A | | MED1 | 0.946964 | 26 | FALSE |
| GSM353617 | LNCaP | POLR2A | | MYC | 1.023162 | 27 | FALSE |
| GSM353650 | RWPE1 | ERG | | RNF2 | 1.476786 | 28 | FALSE |
| GSM353649 | RWPE1 | ERG | | TRIM24 | 2.181727 | 29 | FALSE |
| GSM696842 | VCaP | AR | | ESR1 | 2.360895 | 30 | FALSE |
| GSM696841 | VCaP | AR | | H2AZ | 2.384988 | 31 | FALSE |
| GSM353646 | VCaP | AR | | BRD4 | 3.114181 | 32 | FALSE |
| GSM353638 | VCaP | AR | | RUNX1 | 3.161102 | 33 | FALSE |
| GSM353636 | VCaP | AR | | HDAC1 | 3.173736 | 34 | FALSE |
| GSM353647 | VCaP | ERG | | RELA | 3.44426 | 35 | FALSE |

| | | |
|---|---|---|
| GSM353639 | VCaP | ERG |
| GSM353637 | VCaP | ERG |
| GSM353630 | VCaP | FOXA1 |
| GSM353623 | VCaP | POLR2A |
| GSM353608 | VCaP | POLR2A |
| GSM353607 | VCaP | POLR2A |
| GSM353651 | None | AR |
| GSM353652 | None | ERG |
| GSM353656 | None | POLR2A |
| GSM838400 | C4-2B | RUNX2 |
| GSM801011 | VCaP | AR |
| GSM801012 | VCS2 | AR |
| GSM699632 | LNCaP | NKX3-1 |
| GSM699633 | LNCaP | NKX3-1 |
| GSM699634 | LNCaP | FOXA1 |
| GSM699635 | LNCaP | FOXA1 |
| GSM738817 | PC-3 | ETV4 |
| GSM738818 | PC-3 | JUND |
| GSM738821 | RWPE1 | ETS1 |
| GSM738822 | RWPE1 | GABPA |
| GSM738823 | RWPE1 | ERG |
| GSM738824 | RWPE1 | ETV1 |
| GSM717392 | VCaP | AR |
| GSM717393 | VCaP | AR |
| GSM717394 | VCaP | AR |
| GSM717395 | VCaP | ERG |
| GSM717396 | VCaP | ERG |
| GSM717397 | VCaP | ERG |
| GSM717398 | VCaP | HDAC1 |
| GSM717399 | VCaP | HDAC1 |
| GSM717400 | VCaP | HDAC2 |
| GSM717401 | VCaP | HDAC2 |
| GSM717402 | VCaP | HDAC3 |
| GSM717403 | VCaP | HDAC3 |
| GSM717404 | VCaP | EZH2 |
| GSM717405 | VCaP | EZH2 |
| GSM775364 | LNCaP | AR |
| GSM775365 | LNCaP | AR |
| GSM927071 | RWPE1 | ERG |
| GSM1059397 | LNCaP | POLR2A |
| GSM1059394 | LNCaP | POLR2A |
| GSM1059396 | LNCaP | POLR2A |

| | | | |
|---|---|---|---|
| BRD3 | 3.914287 | 36 | FALSE |
| BRD2 | 4.181282 | 37 | FALSE |
| EZH2 | 4.554295 | 38 | FALSE |
| ERG | 5.898607 | 39 | FALSE |
| DAXX | 7.318007 | 40 | FALSE |
| POLR2A | 7.491274 | 41 | FALSE |
| GABPA | 7.631724 | 42 | FALSE |
| SRC | 11.44643 | 43 | FALSE |
| ETV1 | 12.23014 | 44 | FALSE |
| MRE11A | 13.28621 | 45 | FALSE |
| ARID1A | 19.56408 | 46 | FALSE |
| ERF | 20.03243 | 47 | TRUE |
| CREB1 | 27.94717 | 48 | TRUE |
| NKX3-1 | 39.18543 | 49 | TRUE |
| GRHL2 | 46.4107 | 50 | TRUE |
| HDAC2 | 59.70467 | 51 | TRUE |
| HDAC3 | 133.1866 | 52 | TRUE |
| GATA2 | 148.4117 | 53 | TRUE |
| AR | 149.3384 | 54 | TRUE |
| HOXB13 | 173.5875 | 55 | TRUE |
| PIAS1 | 202.5997 | 56 | TRUE |
| TLE3 | 203.3019 | 57 | TRUE |
| SUMO2 | 272.1432 | 58 | TRUE |
| ASH2L | 278.3605 | 59 | TRUE |
| FOXA1 | 320.1546 | 60 | TRUE |
| HNF4G | 331.7467 | 61 | TRUE |
| TOP1 | 428.4343 | 62 | TRUE |
| NR3C1 | 460.1611 | 63 | TRUE |

| | | |
|---|---|---|
| GSM1059398 | LNCaP | POLR2A |
| GSM1059395 | LNCaP | POLR2A |
| GSM1059393 | LNCaP | POLR2A |
| GSM916524 | LNCaP | FOXA1 |
| GSM916523 | LNCaP | FOXA1 |
| GSM916521 | LNCaP | AR |
| GSM916522 | LNCaP | AR |
| GSM969562 | LNCaP-abl | EZH2 |
| GSM969570 | LNCaP | EZH2 |
| GSM969565 | LNCaP-abl | AR |
| GSM969566 | LNCaP-abl | POLR2A |
| GSM969572 | LNCaP | SUZ12 |
| GSM969564 | LNCaP-abl | SUZ12 |
| GSM984381 | C4-2B | AR |
| GSM984390 | 22RV1 | AR |
| GSM984385 | C4-2B | AR |
| GSM984392 | C4-2B | FOXA1 |
| GSM984386 | LNCaP | AR |
| GSM984389 | 22RV1 | AR |
| GSM984388 | LNCaP | AR |
| GSM984391 | C4-2B | FOXA1 |
| GSM984382 | C4-2B | AR |
| GSM984383 | C4-2B | AR |
| GSM984387 | LNCaP | AR |
| GSM984380 | C4-2B | AR |
| GSM714610 | LNCaP | REPIN1 |
| GSM989640 | LNCaP | NKX3-1 |
| GSM980649 | LNCaP | AR |
| GSM980654 | LNCaP | POLR2A |
| GSM980660 | VCaP | NR3C1 |
| GSM980648 | LNCaP | AR |
| GSM980664 | LNCaP | NR3C1 |
| GSM980658 | VCaP | AR |
| GSM980653 | LNCaP | POLR2A |
| GSM980663 | LNCaP | AR |
| GSM980650 | LNCaP | AR |
| GSM980657 | VCaP | AR |
| GSM980665 | LNCaP | NR3C1 |
| GSM980655 | LNCaP | NR3C1 |
| GSM980651 | LNCaP | AR |
| GSM980647 | LNCaP | AR |

| | | |
|---|---|---|
| GSM980662 | LNCaP | AR |
| GSM980652 | LNCaP | AR |
| GSM1145322 | LNCaP | ETV1 |
| GSM1145324 | LNCaP | AR |
| GSM1145325 | LNCaP | AR |
| GSM947527 | PrEC | CTCF |
| GSM947528 | LNCaP | CTCF |
| GSM923508 | LNCaP | AR |
| GSM923509 | LNCaP | AR |
| GSM1174486 | LNCaP | AR |
| GSM1174482 | LNCaP | AR |
| GSM1174488 | LNCaP | AR |
| GSM1174487 | LNCaP | AR |
| GSM1174481 | LNCaP | AR |
| GSM1174480 | LNCaP | AR |
| GSM1174485 | LNCaP | AR |
| GSM1174491 | LNCaP | AR |
| GSM1174490 | LNCaP | AR |
| GSM1174484 | LNCaP | AR |
| GSM1174483 | LNCaP | AR |
| GSM1045472 | DU145 | EED |
| GSM1045473 | DU145 | EED |
| GSM1056847 | EP156T | TP63 |
| GSM1097496 | LNCaP | SFPQ |
| GSM1097497 | LNCaP | SFPQ |
| GSM1138591 | DU145 | RING1 |
| GSM1138592 | DU145 | RNF2 |
| GSM1138593 | DU145 | RNF2 |
| GSM1138594 | DU145 | BMI1 |
| GSM1138595 | DU145 | BMI1 |
| GSM1249449 | LNCaP | TCF7L2 |
| GSM1274871 | LNCaP | AR |
| GSM1274872 | LNCaP | AR |

| | | |
|---|---|---|
| GSM1274873 | LNCaP | FOXA1 |
| GSM817347 | VCaP | AR |
| GSM817348 | VCaP | AR |
| GSM817349 | VCaP | AR |
| GSM817350 | VCaP | AR |
| GSM817351 | VCaP | AR |
| GSM817352 | VCaP | AR |
| GSM817353 | VCaP | AR |
| GSM817354 | VCaP | AR |
| GSM995943 | PC-3 | AGO1 |
| GSM995944 | PC-3 | AGO2 |
| GSM1207887 | LNCaP | AR |
| GSM1207888 | LNCaP | AR |
| GSM1207889 | LNCaP | E2F1 |
| GSM1207890 | LNCaP | E2F1 |
| GSM1207891 | LNCaP | AR |
| GSM1207892 | LNCaP | AR |
| GSM1207893 | LNCaP | E2F1 |
| GSM1207894 | LNCaP | E2F1 |
| GSM1207895 | LNCaP | AR |
| GSM1207896 | LNCaP | AR |
| GSM1207897 | LNCaP | E2F1 |
| GSM1207898 | LNCaP | E2F1 |
| GSM1207899 | LNCaP | AR |
| GSM1207900 | LNCaP | AR |
| GSM1207901 | LNCaP | E2F1 |
| GSM1207902 | LNCaP | E2F1 |
| GSM1207903 | LNCaP | AR |
| GSM1207904 | LNCaP | AR |
| GSM1207905 | LNCaP | E2F1 |
| GSM1207906 | LNCaP | E2F1 |
| GSM1207907 | LNCaP | AR |
| GSM1207908 | LNCaP | AR |
| GSM1207909 | LNCaP | E2F1 |

| | | |
|---|---|---|
| GSM1207910 | LNCaP | E2F1 |
| GSM1207911 | LNCaP | AR |
| GSM1207912 | LNCaP | AR |
| GSM1207913 | LNCaP | E2F1 |
| GSM1207914 | LNCaP | E2F1 |
| GSM1207915 | LNCaP | AR |
| GSM1207916 | LNCaP | E2F1 |
| GSM1207917 | LNCaP | E2F1 |
| GSM1328945 | VCaP | AR |
| GSM1328947 | VCaP | AR |
| GSM1328950 | VCaP | AR |
| GSM1328952 | VCaP | AR |
| GSM1328953 | VCaP | AR |
| GSM1328954 | VCaP | AR |
| GSM1328955 | VCaP | AR |
| GSM1328956 | VCaP | AR |
| GSM1328957 | VCaP | AR |
| GSM1328958 | VCaP | AR |
| GSM1328959 | VCaP | BRD4 |
| GSM1328960 | VCaP | BRD4 |
| GSM1328961 | VCaP | BRD4 |
| GSM1328962 | VCaP | BRD4 |
| GSM1328963 | VCaP | BRD4 |
| GSM1328964 | VCaP | POLR2A |
| GSM1328965 | VCaP | POLR2A |
| GSM1328966 | VCaP | POLR2A |
| GSM1328967 | VCaP | POLR2A |
| GSM1328968 | VCaP | POLR2A |
| GSM1328969 | VCaP | BRD2 |
| GSM1328970 | VCaP | BRD2 |
| GSM1328971 | VCaP | BRD2 |

| | | |
|---|---|---|
| GSM1328972 | VCaP | BRD3 |
| GSM1328973 | VCaP | BRD3 |
| GSM1328974 | VCaP | BRD3 |
| GSM1328975 | VCaP | BRD4 |
| GSM1328976 | VCaP | BRD4 |
| GSM1328977 | VCaP | BRD4 |
| GSM1328978 | VCaP | ERG |
| GSM1328979 | VCaP | ERG |
| GSM1328980 | VCaP | ERG |
| GSM1328981 | VCaP | ERG |
| GSM1006887 | LNCaP | CTCF |
| GSM1006874 | LNCaP | CTCF |
| GSM1164141 | DU145 | AR |
| GSM1164142 | DU145 | FOXA1 |
| GSM1164143 | DU145 | FOXA1 |
| GSM1164144 | DU145 | FOXA1 |
| GSM1164145 | DU145 | FOXA1 |
| GSM1164146 | DU145 | FOXA1 |
| GSM1164147 | DU145 | FOXA1 |
| GSM1164148 | DU145 | FOXA1 |
| GSM1164149 | DU145 | AR |
| GSM1164150 | DU145 | FOXA1 |
| GSM1164151 | DU145 | AR |
| GSM1164152 | DU145 | FOXA1 |
| GSM1164153 | DU145 | AR |
| GSM1164154 | DU145 | FOXA1 |
| GSM1193656 | VCaP | ERG |
| GSM1193657 | VCaP | ERG |
| GSM1193658 | VCaP | ERG |
| GSM1193659 | VCaP | GABPA |
| GSM1193660 | VCaP | GABPA |

| | | |
|---|---|---|
| GSM1193661 | VCaP | GABPA |
| GSM1308235 | PC-3 | AR |
| GSM1308236 | PC-3 | AR |
| GSM1308237 | PC-3 | AR |
| GSM1308239 | PC-3 | AR |
| GSM1308240 | PC-3 | AR |
| GSM1308241 | PC-3 | AR |
| GSM1327871 | LNCaP | AR |
| GSM1327872 | LNCaP | AR |
| GSM1333368 | LNCaP | WDR5 |
| GSM1333369 | LNCaP | WDR5 |
| GSM1383869 | PrEC | POLR2A |
| GSM1383875 | PC-3 | CBX8 |
| GSM1383877 | PC-3 | CTCF |
| GSM1412317 | 22RV1 | AR |
| GSM1412319 | 22RV1 | AR |
| GSM1415123 | LNCaP | CSNK2A1 |
| GSM1415124 | LNCaP | POLR2A |
| GSM1415125 | LNCaP | POLR2A |
| GSM1422482 | VCaP | ERG |
| GSM1422483 | VCaP | ERG |
| GSM1424526 | DU145 | ETS1 |
| GSM1424527 | DU145 | ELF1 |
| GSM1424528 | DU145 | ELK4 |
| GSM1424529 | DU145 | GABPA |
| GSM1424530 | DU145 | JUND |
| GSM1068136 | LNCaP | FOXA1 |
| GSM1068137 | LNCaP | FOXA1 |
| GSM1069669 | LNCaP | AR |
| GSM1069670 | LNCaP | AR |
| GSM1069681 | LNCaP | AR |

| | | |
|---|---|---|
| GSM1069682 | LNCaP | AR |
| GSM1071276 | LNCaP | AR |
| GSM1071277 | LNCaP | AR |
| GSM1071280 | LNCaP | AR |
| GSM1071281 | LNCaP | AR |
| GSM1071284 | None | AR |
| GSM1071285 | None | AR |
| GSM1071286 | None | AR |
| GSM1071287 | None | AR |
| GSM1071288 | None | AR |
| GSM1071289 | None | AR |
| GSM1071290 | None | AR |
| GSM1071291 | None | AR |
| GSM1071292 | None | AR |
| GSM1071293 | None | AR |
| GSM1071294 | None | AR |
| GSM1071295 | None | AR |
| GSM1076109 | VCaP | ESR1 |
| GSM1076110 | VCaP | ESR1 |
| GSM1076111 | VCaP | ESR1 |
| GSM1076112 | NCI-H660 | ESR1 |
| GSM1236922 | LNCaP | AR |
| GSM1236926 | LNCaP | AR |
| GSM1279769 | LNCaP | KDM1A |
| GSM1279770 | VCaP | KDM1A |
| GSM1340263 | None | PR |
| GSM1410762 | LNCaP | CTBP1 |
| GSM1410763 | LNCaP | CTBP2 |
| GSM1410764 | LNCaP | CTBP1 |
| GSM1410765 | LNCaP | CTBP2 |
| GSM1410768 | VCaP | AR |

| | | |
|---|---|---|
| GSM1410770 | VCaP | FOXP1 |
| GSM1410771 | VCaP | RUNX1 |
| GSM1410772 | VCaP | AR |
| GSM1410773 | VCaP | AR |
| GSM1410774 | VCaP | AR |
| GSM1410775 | VCaP | FOXA1 |
| GSM1410776 | VCaP | FOXA1 |
| GSM1410782 | LNCaP | AR |
| GSM1410784 | VCaP | AR |
| GSM1410785 | VCaP | AR |
| GSM1410787 | LNCaP | AR |
| GSM1410788 | LNCaP | FOXA1 |
| GSM1410789 | LNCaP | FOXA1 |
| GSM1410790 | LNCaP | AR |
| GSM1354830 | VCaP | AR |
| GSM1354831 | VCaP | AR |
| GSM1354832 | VCaP | PIAS1 |
| GSM1354833 | VCaP | PIAS1 |
| GSM1354834 | VCaP | PIAS1 |
| GSM1354835 | VCaP | PIAS1 |
| GSM1354836 | VCaP | FOXA1 |
| GSM1354837 | VCaP | FOXA1 |
| GSM1354838 | VCaP | FOXA1 |
| GSM1354839 | VCaP | FOXA1 |
| GSM1463459 | VCaP | AR |
| GSM1463460 | VCaP | AR |
| GSM1463461 | VCaP | FOXA1 |
| GSM1463462 | VCaP | FOXA1 |
| GSM1463463 | VCaP | FOXA1 |
| GSM1463464 | VCaP | FOXA1 |
| GSM1463465 | VCaP | POLR2A |

| | | |
|---|---|---|
| GSM1463466 | VCaP | POLR2A |
| GSM1489926 | VCaP | ASH2L |
| GSM1489927 | VCaP | ASH2L |
| GSM1527822 | LNCaP | AR |
| GSM1527823 | LNCaP | AR |
| GSM1527834 | LNCaP | AR |
| GSM1527835 | LNCaP | AR |
| GSM1527836 | LNCaP | FOXP1 |
| GSM1527837 | LNCaP | FOXP1 |
| GSM1527839 | LNCaP | RUNX1 |
| GSM1527840 | LNCaP | RUNX1 |
| GSM1527841 | LNCaP | EZH2 |
| GSM1527843 | LNCaP | AR |
| GSM1527844 | LNCaP | AR |
| GSM1543774 | LNCaP | AR |
| GSM1543775 | LNCaP | MRE11A |
| GSM1543776 | LNCaP | MRE11A |
| GSM1543791 | LNCaP | TOP1 |
| GSM1543792 | LNCaP | TOP1 |
| GSM1555570 | VCaP | SUMO2 |
| GSM1555571 | VCaP | SUMO2 |
| GSM1555572 | VCaP | SUMO2 |
| GSM1555573 | VCaP | SUMO2 |
| GSM1600544 | LNCaP | GATA2 |
| GSM1515520 | R1-AD1 | AR |
| GSM1515521 | R1-AD1 | AR |
| GSM1515522 | R1-AD1 | AR |
| GSM1515523 | R1-AD1 | AR |
| GSM1515524 | R1-AD1 | AR |
| GSM1515525 | R1-AD1 | AR |
| GSM1515526 | R1-D567 | AR |

| | | |
|---|---|---|
| GSM1515527 | R1-D567 | AR |
| GSM1515528 | R1-D567 | AR |
| GSM1656410 | LNCaP-abl | E2F1 |
| GSM1385219 | PC-3 | ALKBH3 |
| GSM1678029 | PC-3 | DAXX |
| GSM1678030 | PC-3 | DAXX |
| GSM1678031 | PC-3 | DNMT1 |
| GSM1678032 | PC-3 | DNMT1 |
| GSM941194 | LNCaP | GATA2 |
| GSM941195 | LNCaP | GATA2 |
| GSM1576447 | LNCaP | AR |
| GSM1576449 | LNCaP | VDR |
| GSM1576451 | LNCaP | FOXA1 |
| GSM1613310 | LNCaP | AR |
| GSM1613312 | BicR | AR |
| GSM1613316 | BicR | AR |
| GSM1613320 | BicR | AR |
| GSM1613322 | LNCaP | TET2 |
| GSM1622624 | VCaP | SUMO2 |
| GSM1622625 | VCaP | SUMO2 |
| GSM1622626 | VCaP | SUMO2 |
| GSM1622627 | VCaP | SUMO2 |
| GSM1622628 | VCaP | PIAS1 |
| GSM1622629 | VCaP | PIAS1 |
| GSM1622630 | VCaP | PIAS1 |
| GSM1622631 | VCaP | PIAS1 |
| GSM1622632 | VCaP | POLR2A |
| GSM1622633 | VCaP | POLR2A |
| GSM1622634 | VCaP | POLR2A |
| GSM1622635 | VCaP | POLR2A |
| GSM1622636 | VCaP | POLR2A |
| GSM1622637 | VCaP | POLR2A |

| | | |
|---|---|---|
| GSM1777588 | VCaP | POLR2A |
| GSM1777589 | VCaP | POLR2A |
| GSM1777590 | VCaP | POLR2A |
| GSM1777591 | VCaP | POLR2A |
| GSM1777593 | VCaP | POLR2A |
| GSM1777594 | VCaP | POLR2A |
| GSM1777595 | VCaP | POLR2A |
| GSM1777597 | VCaP | SUMO2 |
| GSM1777598 | VCaP | SUMO2 |
| GSM1777599 | VCaP | SUMO2 |
| GSM1777601 | VCaP | SUMO2 |
| GSM1777604 | VCaP | SUMO2 |
| GSM1777605 | VCaP | SUMO2 |
| GSM1777606 | VCaP | SUMO2 |
| GSM1777608 | VCaP | SUMO2 |
| GSM1777609 | VCaP | SUMO2 |
| GSM1777610 | VCaP | SUMO2 |
| GSM1816381 | DUCaP | AR |
| GSM1816382 | DUCaP | AR |
| GSM1358395 | None | AR |
| GSM1358396 | None | AR |
| GSM1358397 | None | AR |
| GSM1358398 | None | AR |
| GSM1358399 | None | AR |
| GSM1358400 | None | AR |
| GSM1358401 | None | AR |
| GSM1358402 | None | AR |
| GSM1358403 | None | AR |
| GSM1358404 | None | AR |
| GSM1358405 | None | AR |
| GSM1358406 | None | AR |

| | | |
|---|---|---|
| GSM135840<br>7 | None | AR |
| GSM135840<br>8 | None | AR |
| GSM135840<br>9 | None | AR |
| GSM135841<br>0 | None | AR |
| GSM135841<br>1 | None | AR |
| GSM135841<br>2 | None | AR |
| GSM135841<br>3 | None | AR |
| GSM135841<br>4 | None | AR |
| GSM150118<br>4 | LNCaP | AR |
| GSM150118<br>5 | LNCaP | AR |
| GSM150118<br>6 | LNCaP | AR |
| GSM150118<br>7 | LNCaP | AR |
| GSM150118<br>8 | C4-2B | AR |
| GSM150118<br>9 | C4-2B | AR |
| GSM150119<br>0 | C4-2B | AR |
| GSM150119<br>1 | C4-2B | AR |
| GSM152702<br>3 | LNCaP | AR |
| GSM152702<br>4 | LNCaP | AR |
| GSM152702<br>5 | LNCaP | AR |
| GSM152702<br>6 | LNCaP | AR |
| GSM152703<br>3 | CWR22P<br>c | AR |
| GSM152703<br>4 | CWR22P<br>c | AR |
| GSM152703<br>6 | CWR22P<br>c | AR |
| GSM159821<br>8 | None | AR |
| GSM159821<br>9 | None | AR |
| GSM159822<br>0 | None | AR |
| GSM159822<br>1 | None | AR |
| GSM159822<br>2 | None | AR |
| GSM159822<br>3 | None | AR |
| GSM159822<br>4 | None | AR |
| GSM159822<br>5 | None | AR |

| | | |
|---|---|---|
| GSM1716762 | None | FOXA1 |
| GSM1716763 | None | HOXB13 |
| GSM1716764 | LNCaP | HOXB13 |
| GSM1716765 | LHSAR | FOXA1 |
| GSM1716766 | LHSAR | HOXB13 |
| GSM1716767 | LHSAR | HOXB13 |
| GSM1716768 | LHSAR | HOXB13 |
| GSM1716769 | LHSAR | AR |
| GSM1716770 | LHSAR | AR |
| GSM1917726 | PC-3 | EP300 |
| GSM1917727 | PC-3 | EP300 |
| GSM1917728 | PC-3 | EP300 |
| GSM1917729 | PC-3 | EP300 |
| GSM1917730 | PC-3 | EP300 |
| GSM1573653 | LNCaP | CHD1 |
| GSM1573654 | LNCaP | CHD1 |
| GSM1573655 | LNCaP | KDM1A |
| GSM1573656 | LNCaP | KDM1A |
| GSM1679092 | LNCaP-abl | MED1 |
| GSM1679093 | LNCaP-abl | MED1 |
| GSM1679094 | LNCaP-abl | POLR2A |
| GSM1679095 | LNCaP-abl | POLR2A |
| GSM1679096 | LNCaP-abl | MED1 |
| GSM1679097 | LNCaP-abl | POLR2A |
| GSM1679098 | None | MED1 |
| GSM1679110 | LNCaP-abl | MED1 |
| GSM1679111 | LNCaP-abl | MED1 |
| GSM1679112 | LNCaP-abl | POLR2A |
| GSM1679113 | LNCaP-abl | POLR2A |
| GSM1679114 | LNCaP-abl | PAF1 |
| GSM1679115 | LNCaP-abl | SUPT5H |

| | | |
|---|---|---|
| GSM1679116 | LNCaP-abl | MED1 |
| GSM1679118 | LNCaP-abl | MED1 |
| GSM1679119 | LNCaP-abl | MED1 |
| GSM1679120 | LNCaP-abl | MED1 |
| GSM1679124 | LNCaP-abl | MED1 |
| GSM1679125 | LNCaP-abl | MED1 |
| GSM1679126 | LNCaP-abl | MED1 |
| GSM1679127 | LNCaP-abl | POLR2A |
| GSM1679128 | LNCaP-abl | POLR2A |
| GSM1679129 | LNCaP-abl | POLR2A |
| GSM1679130 | LNCaP-abl | POLR2A |
| GSM1679131 | LNCaP-abl | POLR2A |
| GSM1679132 | LNCaP-abl | POLR2A |
| GSM1679133 | LNCaP-abl | POLR2A |
| GSM1679134 | LNCaP-abl | POLR2A |
| GSM1679136 | LNCaP-abl | POLR2A |
| GSM1824834 | LNCaP | CHD1 |
| GSM1824836 | LNCaP | KDM1A |
| GSM1824838 | LNCaP | KDM1A |
| GSM1847365 | LNCaP-abl | POLR2A |
| GSM1847366 | LNCaP-abl | POLR2A |
| GSM1847367 | LNCaP-abl | POLR2A |
| GSM1847368 | LNCaP-abl | POLR2A |
| GSM1847369 | LNCaP-abl | POLR2A |
| GSM1847370 | LNCaP-abl | MED1 |
| GSM1847371 | LNCaP-abl | MED1 |
| GSM1847372 | LNCaP-abl | MED1 |
| GSM1847373 | LNCaP-abl | MED1 |
| GSM1847374 | LNCaP-abl | MED1 |
| GSM1873046 | LNCaP | CREB1 |
| GSM1873045 | LNCaP | CREB1 |

| | | |
|---|---|---|
| GSM1873044 | LNCaP | FOXA1 |
| GSM1873043 | LNCaP | FOXA1 |
| GSM1873042 | LNCaP | CREB1 |
| GSM1873041 | LNCaP | CREB1 |
| GSM1873040 | LNCaP | FOXA1 |
| GSM1873039 | LNCaP | FOXA1 |
| GSM1538431 | LNCaP | CREB1 |
| GSM1538430 | LNCaP | FOXA1 |
| GSM1538429 | LNCaP | CREB1 |
| GSM1691165 | LNCaP | FOXA1 |
| GSM1691164 | LNCaP | FOXA1 |
| GSM1691161 | DU145 | GATA2 |
| GSM1691160 | DU145 | AR |
| GSM1691158 | LNCaP | GATA2 |
| GSM1691157 | LNCaP | FOXA1 |
| GSM1691156 | LNCaP | AR |
| GSM1691155 | LNCaP | GATA2 |
| GSM1691154 | LNCaP | FOXA1 |
| GSM1691153 | LNCaP | AR |
| GSM1691152 | LNCaP | GATA2 |
| GSM1691151 | LNCaP | FOXA1 |
| GSM1691150 | LNCaP | AR |
| GSM1691149 | LNCaP | GATA2 |
| GSM1691148 | LNCaP | FOXA1 |
| GSM1691147 | LNCaP | AR |
| GSM1691146 | LNCaP | GATA2 |
| GSM1691145 | LNCaP | GATA2 |
| GSM1691144 | LNCaP | GATA2 |
| GSM1691143 | LNCaP | GATA2 |
| GSM1691142 | LNCaP | FOXA1 |
| GSM1691141 | LNCaP | AR |

| | | |
|---|---|---|
| GSM1691140 | LNCaP | AR |
| GSM1691139 | LNCaP | AR |
| GSM1691138 | LNCaP | AR |
| GSM1586675 | C4-2 | POLR2A |
| GSM1586674 | C4-2 | POLR2A |
| GSM1586673 | C4-2 | POLR2A |
| GSM1586672 | C4-2 | POLR2A |
| GSM1586665 | LNCaP | AR |
| GSM1586664 | LNCaP | AR |
| GSM1586663 | LNCaP | AR |
| GSM1586662 | LNCaP | AR |
| GSM1586661 | C4-2 | AR |
| GSM1586660 | C4-2 | AR |
| GSM1586659 | C4-2 | AR |
| GSM1586658 | C4-2 | AR |
| GSM1905393 | C4-2 | POLR2A |
| GSM1905392 | C4-2 | POLR2A |
| GSM1905391 | C4-2 | POLR2A |
| GSM1905390 | C4-2 | POLR2A |
| GSM1843416 | C4-2 | POLR2A |
| GSM1843415 | C4-2 | POLR2A |
| GSM1843414 | C4-2 | POLR2A |
| GSM1843413 | C4-2 | POLR2A |
| GSM1843412 | C4-2 | AR |
| GSM1843411 | C4-2 | AR |
| GSM1843410 | C4-2 | AR |
| GSM1843409 | C4-2 | AR |
| GSM2058897 | LNCaP | POU2F1 |
| GSM2058896 | LNCaP | POU2F1 |
| GSM2058894 | LNCaP | AR |
| GSM2058893 | LNCaP | AR |

| | | |
|---|---|---|
| GSM2058892 | LNCaP | FOXA1 |
| GSM2058890 | VCaP | POU2F1 |
| GSM2058889 | VCaP | POU2F1 |
| GSM2058888 | VCaP | POU2F1 |
| GSM2058887 | VCaP | FOXA1 |
| GSM2058886 | VCaP | FOXA1 |
| GSM2058884 | LNCaP | POU2F1 |
| GSM2058883 | LNCaP | POU2F1 |
| GSM2058882 | LNCaP | POU2F1 |
| GSM2058880 | VCaP | AR |
| GSM2058879 | VCaP | AR |
| GSM1868875 | C4-2B | POLR2A |
| GSM1868874 | C4-2B | POLR2A |
| GSM1868867 | C4-2B | AR |
| GSM1868866 | C4-2B | AR |
| GSM1868863 | C4-2B | AR |
| GSM1868862 | C4-2B | AR |
| GSM1697903 | LNCaP | TRIM24 |
| GSM1697902 | LNCaP | TRIM24 |
| GSM1697901 | LNCaP | TRIM24 |
| GSM2086316 | VCaP | ERG |
| GSM2086315 | VCaP | ERG |
| GSM2086314 | VCaP | ERG |
| GSM2086313 | VCaP | ERG |
| GSM2086312 | VCaP | ERG |
| GSM2086311 | VCaP | ERG |
| GSM2086310 | VCaP | ERG |
| GSM2086309 | VCaP | ERG |
| GSM2086308 | VCaP | AR |
| GSM2086307 | VCaP | AR |
| GSM2086306 | VCaP | AR |

| | | |
|---|---|---|
| GSM2086305 | VCaP | AR |
| GSM2086304 | VCaP | AR |
| GSM2086303 | VCaP | AR |
| GSM2086302 | VCaP | AR |
| GSM2086301 | VCaP | AR |
| GSM2086300 | VCaP | AR |
| GSM2086299 | VCaP | AR |
| GSM2086298 | VCaP | AR |
| GSM2086297 | VCaP | AR |
| GSM2086296 | VCaP | AR |
| GSM2086295 | VCaP | AR |
| GSM2086294 | VCaP | AR |
| GSM2086293 | VCaP | AR |
| GSM2086292 | VCaP | AR |
| GSM2086291 | VCaP | AR |
| GSM1863005 | LNCaP | FOXA1 |
| GSM1868502 | LNCaP | CHD4 |
| GSM1868501 | LNCaP | CHD1 |
| GSM1868497 | LNCaP | SMARCA4 |
| GSM1868495 | LNCaP | SMARCA5 |
| GSM1891832 | LNCaP | FOXA1 |
| GSM1891830 | LNCaP | FOXA1 |
| GSM2235693 | VCaP | CTCF |
| GSM2235692 | VCaP | CTCF |
| GSM2235691 | VCaP | CTCF |
| GSM2235690 | VCaP | CTCF |
| GSM2235689 | VCaP | AR |
| GSM2235688 | VCaP | AR |
| GSM2235687 | VCaP | AR |
| GSM2235686 | VCaP | AR |
| GSM2186466 | LNCaP | AR |

| | | |
|---|---|---|
| GSM2186465 | LNCaP | AR |
| GSM2186464 | LNCaP | AR |
| GSM2186463 | LNCaP | AR |
| GSM2186462 | LNCaP | AR |
| GSM2186461 | LNCaP | AR |
| GSM2219885 | LNCaP | RELA |
| GSM2219884 | LNCaP | RELA |
| GSM2219883 | LNCaP | RELA |
| GSM2219882 | LNCaP | RELA |
| GSM2219881 | LNCaP | RELA |
| GSM2219880 | LNCaP | RELA |
| GSM2219879 | LNCaP | RELA |
| GSM2219878 | LNCaP | RELA |
| GSM2219875 | LNCaP | PIAS1 |
| GSM2219874 | LNCaP | PIAS1 |
| GSM2219873 | LNCaP | PIAS1 |
| GSM2219872 | LNCaP | PIAS1 |
| GSM2219871 | LNCaP | PIAS1 |
| GSM2219870 | LNCaP | PIAS1 |
| GSM2219869 | LNCaP | PIAS1 |
| GSM2219868 | LNCaP | PIAS1 |
| GSM2219867 | LNCaP | FOXA1 |
| GSM2219866 | LNCaP | FOXA1 |
| GSM2219865 | LNCaP | FOXA1 |
| GSM2219864 | LNCaP | FOXA1 |
| GSM2219863 | LNCaP | FOXA1 |
| GSM2219862 | LNCaP | FOXA1 |
| GSM2219861 | LNCaP | FOXA1 |
| GSM2219860 | LNCaP | FOXA1 |
| GSM2219859 | LNCaP | AR |
| GSM2219858 | LNCaP | AR |

| | | |
|---|---|---|
| GSM2219857 | LNCaP | AR |
| GSM2219856 | LNCaP | AR |
| GSM2219855 | LNCaP | AR |
| GSM2219854 | LNCaP | AR |
| GSM2219853 | LNCaP | AR |
| GSM2219852 | LNCaP | AR |
| GSM2219851 | LNCaP | RELA |
| GSM2219850 | LNCaP | RELA |
| GSM2219849 | LNCaP | RELA |
| GSM2219848 | LNCaP | RELA |
| GSM1933889 | LNCaP | NANOG |
| GSM1933888 | LNCaP | NANOG |
| GSM1933887 | LNCaP | NANOG |
| GSM1933886 | LNCaP | NANOG |
| GSM1933885 | LNCaP | NANOG |
| GSM1899558 | VCaP | ERG |
| GSM2195114 | VCaP | EWSR1 |
| GSM2195112 | RWPE1 | ERG |
| GSM2195110 | RWPE1 | ERG |
| GSM2195108 | RWPE1 | ERG |
| GSM2195106 | RWPE1 | ERG |
| GSM2195103 | RWPE1 | ERG |
| GSM1868499 | LNCaP | SMARCA2 |
| GSM2024822 | VCaP | SOX9 |
| GSM2427951 | None | AR |
| GSM2427950 | None | AR |
| GSM2427948 | None | SRC |
| GSM2427947 | None | SRC |
| GSM2427945 | None | AR |
| GSM2427944 | None | AR |
| GSM2092845 | LNCaP | AR |

| | | |
|---|---|---|
| GSM2092844 | LNCaP | AR |
| GSM2298952 | DU145 | ETS1 |
| GSM2298951 | DU145 | ETS1 |
| GSM2422849 | None | CHD1 |
| GSM2422848 | None | CHD1 |
| GSM2422845 | None | CHD1 |
| GSM2422844 | None | CHD1 |
| GSM2122805 | LNCaP | AR |
| GSM2122804 | LNCaP | AR |
| GSM2122803 | LNCaP | GRHL2 |
| GSM2122802 | LNCaP | GRHL2 |
| GSM1374122 | PC-3M-luc | KDM1A |
| GSM1907206 | LNCaP | MYC |
| GSM1907205 | LNCaP | MYC |
| GSM1907204 | LNCaP | MYC |
| GSM1907203 | LNCaP | MYC |
| GSM1907202 | LNCaP | AR |
| GSM1907201 | LNCaP | AR |
| GSM1907200 | LNCaP | AR |
| GSM1907199 | LNCaP | AR |
| GSM2412293 | None | AR |
| GSM2412292 | None | AR |
| GSM1586671 | LNCaP | CCNT1 |
| GSM1586670 | LNCaP | CCNT1 |
| GSM1586669 | C4-2 | CCNT1 |
| GSM1586668 | C4-2 | CCNT1 |
| GSM1586667 | C4-2 | CCNT1 |
| GSM1586666 | C4-2 | CCNT1 |
| GSM1868498 | LNCaP | SMARCA1 |
| GSM2187248 | DU145 | ZMYND8 |
| GSM2187246 | DU145 | ZMYND8 |

| | | |
|---|---|---|
| GSM229895<br>0 | DU145 | ZMYND1<br>1 |
| GSM227716<br>0 | 22RV1 | FOXA1 |
| GSM230296<br>8 | LNCaP | AR |
| GSM257912<br>1 | C4-2 | BMI1 |
| GSM257912<br>2 | C4-2 | AR |
| GSM239382<br>9 | LNCaP | AR |
| GSM261244<br>8 | VCaP | ERF |
| GSM261244<br>9 | VCaP | ERF |
| GSM235045<br>4 | C4-2 | BRD4 |
| GSM235045<br>3 | C4-2 | BRD4 |
| GSM235045<br>0 | C4-2 | BRD4 |
| GSM235045<br>1 | C4-2 | BRD4 |
| GSM290953<br>7 | C4-2B | POLR2A |
| GSM290953<br>6 | PrEC | ZFX |
| GSM239384<br>1 | LNCaP | AR |
| GSM239384<br>3 | LNCaP | AR |
| GSM230296<br>5 | LNCaP | AR |
| GSM230296<br>4 | LNCaP | AR |
| GSM230296<br>7 | LNCaP | AR |
| GSM271692<br>1 | LNCaP | AR |
| GSM271692<br>3 | LNCaP | AR |
| GSM272973<br>3 | R1-D567 | AR |
| GSM272973<br>0 | R1-D567 | AR |
| GSM227715<br>3 | LNCaP | HNF4G |
| GSM227715<br>0 | LNCaP | FOXA1 |
| GSM198141<br>3 | PrEC | H2AZ |
| GSM198141<br>5 | LNCaP | H2AZ |
| GSM198141<br>4 | PrEC | H2AZ |
| GSM198141<br>6 | LNCaP | H2AZ |
| GSM212257<br>9 | LNCaP-<br>abl | AR |
| GSM253721<br>5 | 22RV1 | HOXB13 |

| | | |
|---|---|---|
| GSM2537214 | 22RV1 | HOXB13 |
| GSM2537217 | 22RV1 | FOXA1 |
| GSM2277159 | 22RV1 | AR |
| GSM2277158 | 22RV1 | AR |
| GSM2277151 | LNCaP | FOXA1 |
| GSM2277157 | LNCaP | HNF4G |
| GSM2277156 | LNCaP | HNF4G |
| GSM2480812 | LNCaP | FOXA1 |
| GSM2480813 | LNCaP | FOXA1 |
| GSM2480810 | LNCaP | SMARCA4 |
| GSM2480811 | LNCaP | SMARCA4 |
| GSM2480816 | LNCaP | HOXB13 |
| GSM2480817 | LNCaP | HOXB13 |
| GSM2480814 | LNCaP | FOXA1 |
| GSM2480815 | LNCaP | FOXA1 |
| GSM2480818 | LNCaP | HOXB13 |
| GSM2480819 | LNCaP | HOXB13 |
| GSM2537228 | VCaP | FOXA1 |
| GSM2537229 | VCaP | FOXA1 |
| GSM2537225 | VCaP | FOXA1 |
| GSM2537226 | VCaP | FOXA1 |
| GSM2537227 | VCaP | FOXA1 |
| GSM2537216 | 22RV1 | FOXA1 |
| GSM2480830 | LNCaP | WDHD1 |
| GSM2480831 | LNCaP | WDHD1 |
| GSM2480832 | LNCaP | WDHD1 |
| GSM2480833 | LNCaP | WDHD1 |
| GSM2729729 | R1-D567 | POLR2A |
| GSM2741768 | C4-2B | ZFX |
| GSM2393839 | LNCaP | AR |
| GSM2393831 | LNCaP | AR |

| | | |
|---|---|---|
| GSM2393833 | LNCaP | AR |
| GSM2393835 | LNCaP | AR |
| GSM2393837 | LNCaP | AR |
| GSM2612451 | VCaP | ERF |
| GSM2612450 | VCaP | ERF |
| GSM2612457 | VCaP | ERG |
| GSM2612456 | VCaP | AR |
| GSM2350449 | C4-2 | BRD4 |
| GSM2350448 | C4-2 | BRD4 |
| GSM2350445 | C4-2 | BRD4 |
| GSM2350447 | C4-2 | BRD4 |
| GSM2350446 | C4-2 | BRD4 |
| GSM2350452 | C4-2 | BRD4 |
| GSM2716917 | LNCaP | AR |
| GSM2716919 | LNCaP | AR |
| GSM2771596 | LREX' | BRD4 |
| GSM2771597 | LREX' | BRD4 |
| GSM2277161 | 22RV1 | FOXA1 |
| GSM2277166 | 22RV1 | HNF4G |
| GSM2122581 | LNCaP-abl | AR |
| GSM2122580 | LNCaP-abl | AR |
| GSM2277148 | LNCaP | AR |
| GSM2277149 | LNCaP | HNF4G |
| GSM2537218 | LNCaP | HOXB13 |
| GSM2537213 | 22RV1 | AR |
| GSM2537212 | 22RV1 | AR |
| GSM2741769 | C4-2B | ZFX |
| GSM2480801 | LNCaP | AR |
| GSM2480800 | LNCaP | AR |
| GSM2480802 | LNCaP | AR |
| GSM2480805 | LNCaP | ARID1A |

| | | |
|---|---|---|
| GSM2480804 | LNCaP | ARID1A |
| GSM2480807 | LNCaP | ARID1A |
| GSM2480806 | LNCaP | ARID1A |
| GSM2480809 | LNCaP | SMARCA4 |
| GSM2480808 | LNCaP | SMARCA4 |
| GSM2277167 | 22RV1 | HNF4G |
| GSM2537233 | VCaP | HOXB13 |
| GSM2537232 | VCaP | HOXB13 |
| GSM2537231 | VCaP | HOXB13 |
| GSM2537230 | VCaP | FOXA1 |
| GSM2537235 | VCaP | HOXB13 |
| GSM2537234 | VCaP | HOXB13 |
| GSM2480828 | LNCaP | TRIM28 |
| GSM2480823 | LNCaP | TLE3 |
| GSM2480822 | LNCaP | TLE3 |
| GSM2480820 | LNCaP | HOXB13 |
| GSM2480827 | LNCaP | TRIM28 |
| GSM2480826 | LNCaP | TRIM28 |
| GSM2480825 | LNCaP | TLE3 |
| GSM2480824 | LNCaP | TLE3 |
| GSM2463796 | LNCaP | MTOR |
| GSM2305255 | LNCaP | MYC |
| GSM2305254 | LNCaP | MYC |
| GSM2305253 | LNCaP | MYC |
| GSM2305252 | LNCaP | MYC |
| GSM2305251 | LNCaP | MYC |
