## Supplementary Data for "Enhancer profiling identifies epigenetic markers of endocrine resistance and reveals therapeutic options for metastatic castration-resistant prostate cancer patients"

The supplementary file includes 1 supplement with additional clinical methods.

Tesa M. Severson[1,2,3,*], Yanyun Zhu[1,2,*], Stefan Prekovic[1,2,*], Karianne Schuurman[1], Holly M. Nguyen[4], Lisha G. Brown[4], Sini Hakkola[5], Yongsoo Kim[1,6], Jeroen Kneppers[1,2], Simon Linder[1,2], Suzan Stelloo[1,2,7], Cor Lieftink[3], Michiel van der Heijden[3,8], Matti Nykter[5], Vincent van der Noort[9], Joyce Sanders[10], Ben Morris[3], Guido Jenster[11], Geert JLH van Leenders[12], Mark Pomerantz[13], Matthew L. Freedman[13,14], Roderick L. Beijersbergen[6], Alfonso Urbanucci[5,15], Lodewyk Wessels[2,3,16], Eva Corey[4], Wilbert Zwart[1,2,17#], Andries M. Bergman[1,8#]

**1** Division of Oncogenomics, The Netherlands Cancer Institute, Amsterdam, the Netherlands
**2** Oncode Institute, the Netherlands
**3** Division of Molecular Carcinogenesis, The Netherlands Cancer Institute, Amsterdam, the Netherlands
**4** Department of Urology, University of Washington, Seattle, WA, USA
**5** Prostate Cancer Research Center, Faculty of Medicine and Health Technology, Tampere University and Tays Cancer Centre, Tampere, Finland
**6** present working address: Department of Pathology, Cancer Center Amsterdam, Amsterdam UMC, Vrije Universiteit Amsterdam, Amsterdam, the Netherlands
**7** present working address: Department of Molecular Biology, Faculty of Science, Radboud Institute for Molecular Life Sciences, Oncode Institute, Radboud University Nijmegen, 6525 GA Nijmegen, The Netherlands.
**8** Division of Medical Oncology, The Netherlands Cancer Institute, Amsterdam, the Netherlands
**9** Department of Biometrics, The Netherlands Cancer Institute, Amsterdam, The Netherlands.
**10** Department of Pathology, The Netherlands Cancer Institute, Amsterdam, the Netherlands
**11** Department of Urology, Erasmus MC, Rotterdam, The Netherlands
**12** Department of Pathology, Erasmus MC Cancer Institute, University Medical Centre, Rotterdam, The Netherlands.
**13** Department of Medical Oncology, Dana-Farber Cancer Institute, Harvard Medical School, Boston, MA, USA.
**14** The Eli and Edythe L. Broad Institute, Cambridge, MA, USA
**15** Department of Tumor Biology, Institute for Cancer Research, Oslo University Hospital, Oslo, Norway
**16** Department of EEMCS, Delft University of Technology, Delft, The Netherlands.
**17** Laboratory of Chemical Biology and Institute for Complex Molecular Systems, Department of Biomedical Engineering, Eindhoven University of Technology, Eindhoven, The Netherlands.

*shared first authors

#### Supplementary Clinical Methods

*Baseline characteristics and response data*

In Supplementary Table 1, the baseline characteristics of all 64 included patients (whole population) are summarized. Median age of the patients was 69 years and the majority of patients had skeletal (75.0%) and/or lymph node (70.3%) metastases, while 34.4% had visceral organ metastases. Virtually all patients (93.8%) had PSA progression, while radiographic progression was established in 81.3% of patients prior to enzalutamide treatment. Approximately one-third of patients (32.8%) received enzalutamide as a first line mCRPC treatment, while all other patients previously received other treatments, including docetaxel chemotherapy (62.5%) and the AR-signaling targeting agent abiraterone (23.4%). Outcomes of treatment, after a median of 17 (IQR: 9-30) months follow-up, are summarized in Supplementary Table 2. All patients in the trial were treated with enzalutamide (160 mg once daily), except for one patient who was treated with abiraterone instead. The median duration of treatment was 22.7 (IQR: 13-45) weeks and 46.9% of patients had a ≥ 50% decrease of serum PSA from baseline. Complete or partial response was found in 18.8% of patients and 17.2% of patients had radiographic stable disease after 12 weeks of treatment. The primary endpoint of the trial, Time to PSA progression (TTPP) was 17.7 (IQR 12-35) weeks, while median overall survival was 14.5 months.

Rate of PSA response in the current study (enzalutamide-treated in first, second and later treatment lines) was lower than in the randomized controlled trial of first line enzalutamide (PREVAIL trial) (78%)[1], but in line with the randomized controlled trial of second line enzalutamide (AFFIRM trial) (54%)[2]. The primary endpoint of the trial, Time to PSA progression (TTPP) was 17.7 (IQR 12-35) weeks which falls within the expected range considering 23.4% of patients in the current trial were previously treated with abiraterone and consequently likely to have a lower likelihood of durable response due to cross-resistance[3]. However, the composition of patients in the trial, based on the line of treatment, deferred from the assumption that 60% percent of patients would have been treated with enzalutamide as a third or higher line of mCRPC treatment. This means that the trial population had fewer prior treatments than anticipated. The TTPP in the current trial was shorter than in the AFFIRM trial (36 weeks)[2]. Complete or partial response was found in 18.8% of patients and 17.2% of patients had radiographic stable disease after 12 weeks of treatment. Overall survival was 14.5 months which was shorter than the 18.4 months, reported in the AFFIRM study[2].

In conclusion, enzalutamide treatment resulted in a TTPP within the expected range. The TTPP was shorter than in the registration studies PREVAIL and AFFIRM, which is explained by the miscellaneous population with more pretreatments, most notably prior abiraterone.

*Trial procedure and patient outcomes*

Baseline studies included radiographic evaluation (CT scan of thorax and abdomen), physical examination, ECOG performance score, blood cell counts and serum chemistry and a biopsy from a metastatic site. Subsequently, patients were treated with enzalutamide at a starting dose of 160 mg/day (4 tablets of 40 mg once daily). Dose adjustments to as low as 80 mg/day were allowed. During the course of enzalutamide treatment, physical examination, ECOG performance score, blood analysis, including PSA measurements, was performed every four weeks at the outpatient clinic. A radiographic response was evaluated at 12 weeks by means of a CT scan of thorax and abdomen (RECIST1.1). Patients with bone only metastases were considered progressive when new metastases were detected. In these patients, a response to treatment could not be evaluated. Enzalutamide treatment was continued until progression or intolerance and selection of subsequent treatment. Frequency of visits to the outpatient clinic and assessments beyond enzalutamide treatment were at the discretion of the physician. Primary endpoint of the study is Time to PSA Progression (TTPP) defined as time from inclusion into the trial until date of a confirmed second PSA rise (PCWG3). Secondary recorded clinical outcomes are, rate of ≥50% serum PSA decrease from baseline, radiographic response (RECIST 1.1 criteria) and overall survival (OS) defined as time from inclusion into the trial until death.

*Trial statistical analysis*

For biomarker discovery purposes, we aimed to construct a cohort of enzalutamide treated patients at various lines of mCRPC treatment in a Phase 2 clinical trial, which is representative for the general population. The first line enzalutamide PREVAIL study[1] showed an 11.2 month (48.7 weeks) time to PSA progression (TTPP), while the second line enzalutamide AFFIRM study[2] showed an 8.3 month (36.1 weeks) TTPP. A retrospective study into enzalutamide treated patients in fourth or fifth line, suggested a TTPP of 3.6 months (15.7 weeks)[4]. We assumed to enroll 20% of patients treated with enzalutamide as a first line treatment, 20% as a second and 60% of patients in a higher line of treatment. Consequently, we expect a TTPP in this miscellaneous population of 22 weeks. Inclusion of 60 patients will allow the detection of this median TTPP with 95% confidence intervals of 16.4-33.6 weeks. This expected range is sufficiently narrow, that when the true TTPP falls within these limits, it suggests that results of the trial are representative and can be interpreted clinically. All time-to-event endpoints were estimated by the Kaplan-Meier method. Differences in normally distributed baseline characteristics and study outcomes between the whole population and the evaluable pre-treatment biopsy and post-treatment biopsy sub populations were evaluated by a student t-test.

Supplemental Methods References


1 Beer, T. M. *et al.* Enzalutamide in metastatic prostate cancer before chemotherapy. *N Engl J Med* **371**, 424-433 (2014). https://doi.org:10.1056/NEJMoa1405095

2 Scher, H. I. *et al.* Increased survival with enzalutamide in prostate cancer after chemotherapy. *N Engl J Med* **367**, 1187-1197 (2012). https://doi.org:10.1056/NEJMoa1207506

3 Khalaf, D. J. *et al.* Optimal sequencing of enzalutamide and abiraterone acetate plus prednisone in metastatic castration-resistant prostate cancer: a multicentre, randomised, open-label, phase 2, crossover trial. *Lancet Oncol* **20**, 1730-1739 (2019). https://doi.org:10.1016/S1470-2045(19)30688-6

4 Badrising, S. K. *et al.* Enzalutamide as a Fourth- or Fifth-Line Treatment Option for Metastatic Castration-Resistant Prostate Cancer. *Oncology* **91**, 267-273 (2016). https://doi.org:10.1159/000448219
